# OCT2Hist: Non-Invasive Virtual Biopsy Using Optical Coherence Tomography

**DOI:** 10.1101/2021.03.31.21254733

**Authors:** Yonatan Winetraub, Edwin Yuan, Itamar Terem, Caroline Yu, Warren Chan, Hanh Do, Saba Shevidi, Maiya Mao, Jacqueline Yu, Megan Hong, Erick Blankenberg, Kerri E. Rieger, Steven Chu, Sumaira Aasi, Kavita Y. Sarin, Adam de la Zerda

## Abstract

Histological haematoxylin and eosin–stained (H&E) tissue sections are used as the gold standard for pathologic detection of cancer, tumour margin detection, and disease diagnosis^1^. Producing H&E sections, however, is invasive and time-consuming. Non-invasive optical imaging modalities, such as optical coherence tomography (OCT), permit label-free, micron-scale 3D imaging of biological tissue microstructure with significant depth (up to 1mm) and large fields-of-view^2^, but are difficult to interpret and correlate with clinical ground truth without specialized training^3^. Here we introduce the concept of a virtual biopsy, using generative neural networks to synthesize virtual H&E sections from OCT images. To do so we have developed a novel technique, “optical barcoding”, which has allowed us to repeatedly extract the 2D OCT slice from a 3D OCT volume that corresponds to a given H&E tissue section, with very high alignment precision down to 25 microns. Using 1,005 prospectively collected human skin sections from Mohs surgery operations of 71 patients, we constructed the largest dataset of H&E images and their corresponding precisely aligned OCT images, and trained a conditional generative adversarial network^4^ on these image pairs. Our results demonstrate the ability to use OCT images to generate high-fidelity virtual H&E sections and entire 3D H&E volumes. Applying this trained neural network to *in vivo* OCT images should enable physicians to readily incorporate OCT imaging into their clinical practice, reducing the number of unnecessary biopsy procedures.

## Introduction

Histopathology, such as H&E tissue sections, has long been the gold standard for disease diagnosis by clinicians. It provides a view of the tissue down to the micron scale, and is a routine part of diagnostic procedures for both cancer^5^ and non-cancer pathologies^6^, as well as surgical procedures such as intraoperative frozen section analysis and Mohs surgery^7^. However, the current histopathology process is an invasive and time consuming task that can take anywhere from a few hours to several days due to its multiple processing steps, including tissue incision, formalin fixation, paraffin embedding, tissue sectioning and staining^1^ (Fig. 1a).

**Figure 1:**
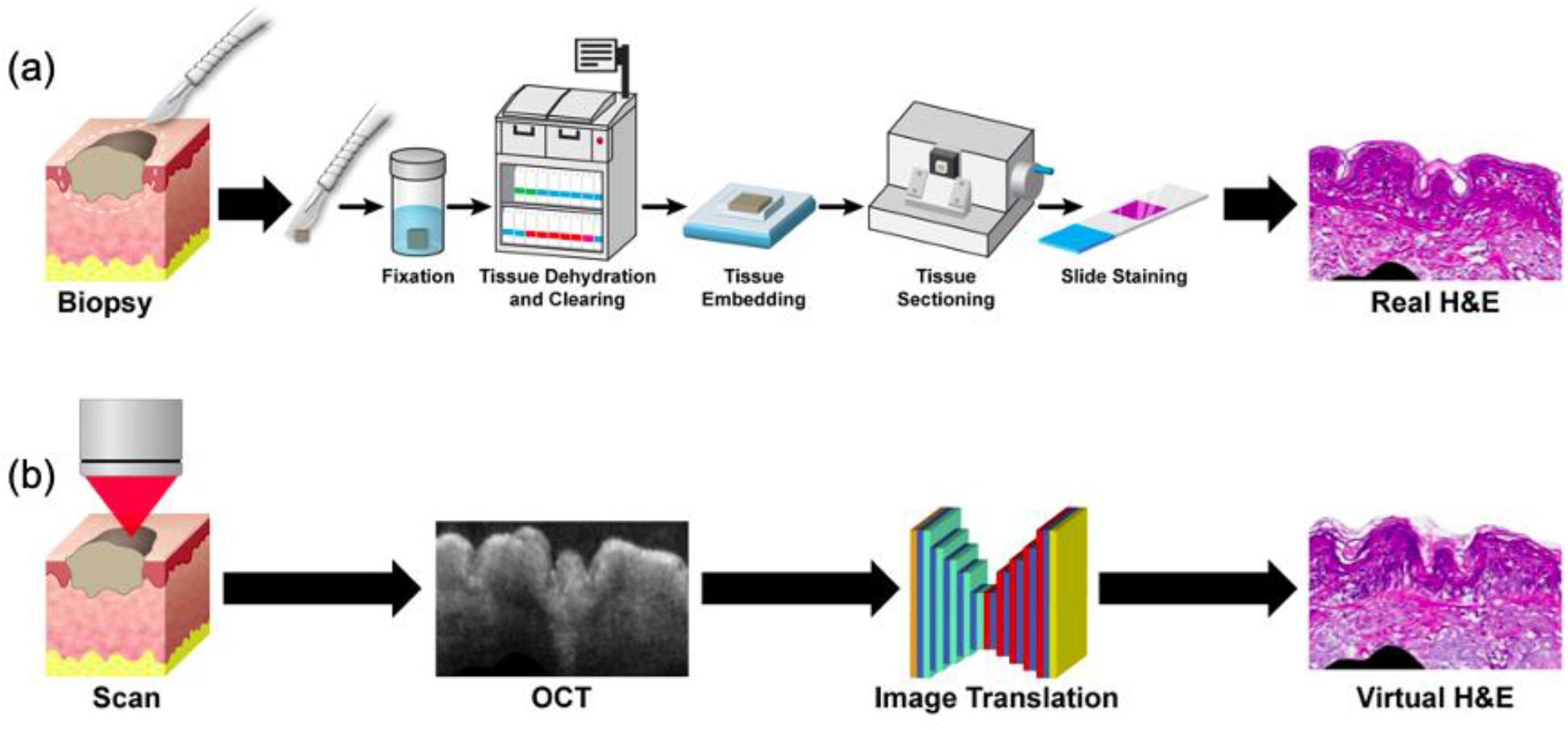
Traditional biopsy versus virtual biopsy. (a) In a traditional biopsy, tissue is first excised, and then undergoes multiple steps including fixation, dehydration, clearing, embedding, sectioning and staining to yield 2D H&E sections which can be examined under a microscope. (b) For a virtual biopsy, an OCT scan of tissue is acquired, and a trained neural network: OCT2Hist transforms the 2D OCT image into a corresponding H&E-like image.

A potential alternative to histopathology is biomedical imaging. In particular, optical imaging modalities are among the highest resolution methods for in vivo imaging^8,9^. In recent years, clinical applications have emerged such as fluorescence-guided surgery^10^, autofluorescence imaging for virtual histology staining^11^, nonlinear microscopy methods for stain-free histopathology^12^, stimulated Raman histology for intraoperative brain tumour diagnosis^13^, hyperspectral imaging for cancer margin detection^14^ and OCT for skin cancer diagnosis^15^. In spite of the high resolution, however, many types of optical image remain challenging for clinical professionals to interpret.

This problem is especially evident for coherent modalities, such as OCT, which frequently suffer from poor tissue contrast due to speckle noise^16^. Studies of OCT have found high sensitivity and specificity for diagnosing certain dermatological pathologies, such as Basal Cell Carcinoma, based on OCT images^17–19^—but only after the clinicians received extensive training in interpreting images. Despite OCT’s demonstrated suitability for use with epithelial cancers such as skin cancer, it has found only a limited clinical role as a diagnostic tool for these cancers in practice^20,21^.

In this work, we introduce a novel 3D virtual biopsy technology (“OCT2Hist”), designed to circumvent the complications of traditional biopsy by using a generative neural network^4^ to transform OCT images into H&E histology-like images (Fig. 1). This approach offers many significant advantages: OCT is quick and entirely non-invasive, acquiring a single 2D image within milliseconds^15^. It requires neither the lengthy tissue preparation procedures nor the significant material and personnel resources needed for traditional biopsy. Furthermore, by non-invasively acquiring multiple images over time, the technique would be far more suitable for tracking the development of a disease *in vivo*.

However, image-to-image translation with neural networks has stringent data requirements for a task to be learned successfully. Ideally, the training examples should be well-aligned to facilitate learning. In our case, each OCT image must be registered to its corresponding H&E image with the highest precision possible^4^. High resolution registration of medical images with H&E images requires a precise match in 9 degrees of freedom (3 translations, 3-axis rotations, and shrinkage/scale change in all axis) and had not previously been adequately achieved^22^, and certainly not in a systematic and repeatable manner^22^. Previous work relied on tissue landmarks, careful orientation of tissue cutting, and substantial luck to arrive at an acceptable OCT-H&E image pair.^23^ Even in the best-case scenario, it was difficult to correlate tissue features in the OCT with those in the H&E image with high precision. We estimate the registration precision achieved using these methods was 500 microns at best.

## Results

To overcome this registration issue, we developed a novel method of “optical barcoding” that allows us to compute the 2D plane within a 3D OCT volume that corresponds to a particular 2D histology image (Fig. 2). With this technique we assembled a database of 1,005 aligned OCT– H&E image pairs of human skin samples. The precision of alignment was better than 25 microns, about an order of magnitude better than the state of the art. These image pairs were aligned with sufficient precision to train a conditional generative adversarial network (cGAN) to perform image-to-image translation from OCT images to H&E-like images. Tests of this translation process on OCT images of new skin samples showed a very high level of agreement between our generated H&E-like images and the corresponding actual H&E sections of the new samples. Our OCT2Hist system has thereby demonstrated a proof-of-principle of the virtual biopsy concept.

**Figure 2:**
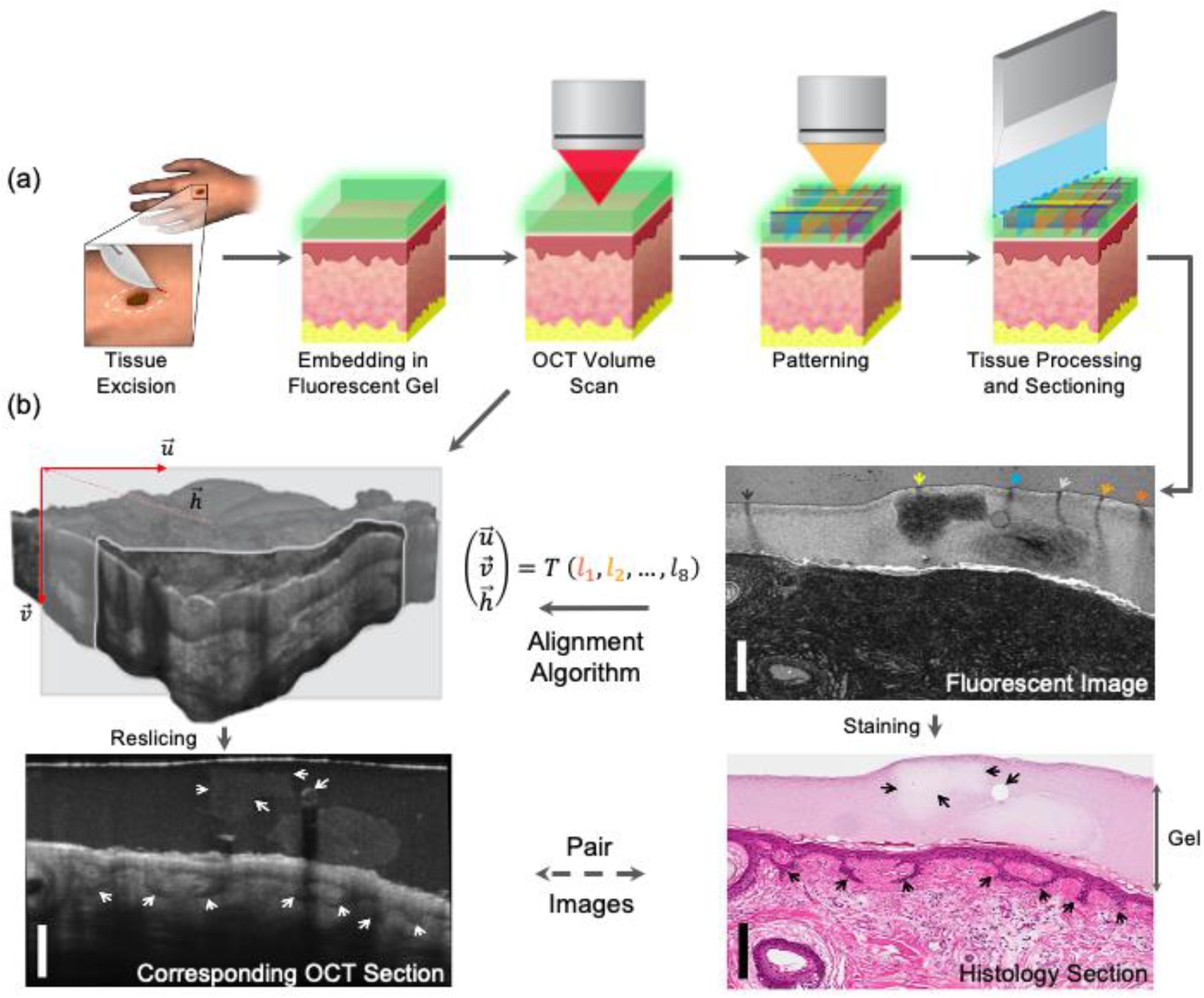
Illustration of the optical barcoding method used to create precisely aligned OCT-H&E image pairs from ex vivo tissue samples. (a) Top row, left to right: The collected tissue specimen is encased in the fluorescent gel, after an OCT volume scan is taken and a diode laser patterns the optical barcode by photobleaching the fluorescent gel, then the tissue undergoes histological processing and tissue sectioning. (b) The tissue section is imaged (top right) to produce a 2D fluorescence image containing the optical barcode (denoted by the colored arrows), and then H&E stain (bottom right). The barcode is used to reslice the 3D OCT volume (top left) and to extract a 2D OCT image (bottom left) that physically corresponds to the H&E section (bottom right). The black and white arrows pointing to strands of epithelial cells in the epithelium as well as a gelatin chunk in the tissue-encasing gel in both H&E and its corresponding OCT images).

### Alignment using optical barcoding

Our optical barcoding method relies on encasing a freshly excised tissue sample within a transparent fluorescent agar-gelatin gel (Fig. 2; see also Methods section below and Supplementary Materials). We acquire a 3D OCT volume image of the tissue within the gel, and then a specific geometric pattern is photobleached into the gel by a second laser wavelength sent through the OCT optical system (Fig 2a). This pattern forms a barcode marker that is preserved through the standard histological process and encodes all orientation, position and scaling information about how the section was cut (Fig 2b). An alignment algorithm uses the appearance of this marker in the histology image to compute a 2D reslice of the OCT volume that corresponds in one-to-one fashion with the histology image. Our technique is repeatable and we regularly achieve registration precision down to 25 microns, as verified by bead-in-gel experiments (Fig. S7) and by correlating features between the OCT and histology images (Fig. S8).

We applied this barcoding technique to histology sections from 119 human skin samples freshly excised during Mohs surgery (IRB-24307), thereby assembling a large dataset of aligned OCT– H&E image pairs. In addition to the alignment computed from the optical barcode, we manually performed additional fine alignment of the OCT-H&E image pairs based on tissue features within both images, to achieve the highest quality of alignment possible. We removed samples with low-quality alignment, or low quality of either OCT or H&E images, and arrived at a dataset of 1,005 aligned OCT-H&E image pairs (one image pair per histology section) taken from 71 of the skin samples. The dataset was subject to further digital processing for stain color normalization and removal of low-SNR image regions, before the images were used for the machine learning task.

### Machine learning

To produce our OCT2Hist image translator, we developed a conditional GAN model based on the pix2pix framework, but with some changes which were found to improve the quality of the generated images. We trained our cGAN model^4^ on the dataset of 553 high-quality OCT-H&E image pairs from 38 patients (54% of dataset). We then tested its performance on patient samples that it had not encountered before (452 high-quality OCT-H&E image pairs from the other 33 patients). Our training data included a large number of skin samples from the scalp and regions of the face and neck, and smaller numbers from the legs and chest (Fig. 3a, S14, S15).

**Figure 3:**
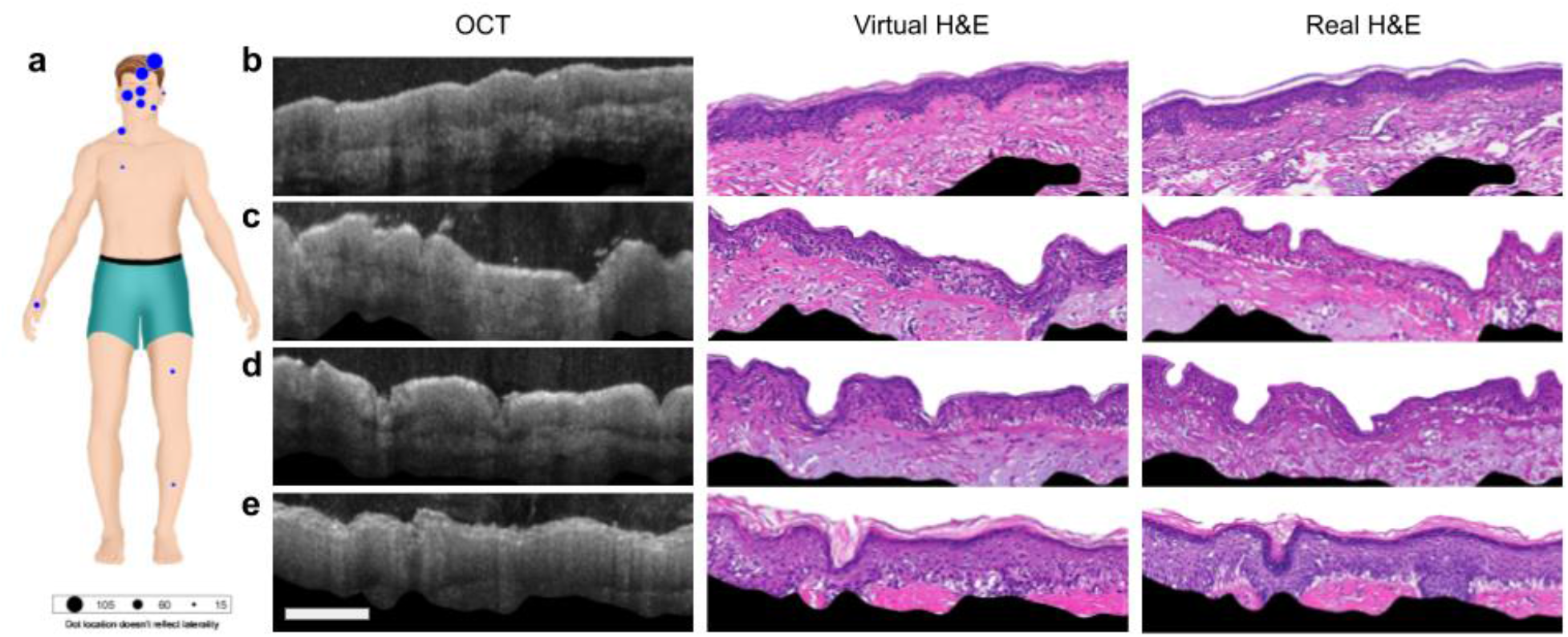
Generating virtual histology H&E images from OCT. (a) Locations of biopsies used in our training set (blue dots). Panel at right shows examples (from the testing set) of an OCT image, the computer generated virtual H&E image, and the corresponding actual histology image from face region where we had a dense training set (b,c,d), and from the forearm (e) where we had no nearby training set. Features as small as tens of microns in size, as found in the shape of the epithelium, are matched. We do not expect smaller features to match as our OCT scanner does not have the resolution or information density to accurately generate smaller features. Scale bar is 200 microns.

Figure 3b–d shows example test results of OCT images from the face and forearm, each one alongside the virtual H&E generated by the neural network, and the real H&E that the OCT slice had been aligned to by our barcoding method. In the OCT images, the nuclei-rich epithelial layer appears darker, while the highly scattering connective tissue of the stroma tends to appear brighter. The generated H&E images demonstrate the neural network’s understanding of these features.

The first sample from the face (Fig. 3b) has a well-defined epithelial layer and strong signal and contrast within the stroma. Notice that the epithelium shrinks during histological processing, and the image translator takes this into account, accurately reproducing the epithelial thickness of the H&E images. Part of a hair follicle is also visible on the left side of the H&E image. The follicle is less apparent in the virtual H&E image, possibly due to imperfect alignment or difficulty seeing it in the OCT image. The final sample (Fig. 3d) is of arm skin with ridge-like epithelium.

Interestingly, the neural network generated a high quality H&E virtual image despite seeing no examples of arm skin in its training set. It is also recognized from the OCT image that this sample has characteristics of sun damage, as seen in discolored, lighter gray tissue found throughout the stroma of both the virtual and ground truth H&E. Most test set samples show similar results (Fig. S17).

To further test the quality of the virtual H&E images, we randomly selected 70 H&E images (35 real, 35 virtual) from different patients in the test set, and gave them to two pathologists to assess in a double-blind experiment. The pathologists correctly determined whether a biopsy image was real or virtual only 69% of the time, indicating that the virtual images are not easily distinguishable from real ones. We believe that by further increasing the training set, we can further improve neural network performances (see Supplementary Materials).

By generating virtual H&E images slice-by-slice, we can also create an entire virtual H&E volume from a corresponding OCT volume (Fig. 4). The virtual H&E volume can be resliced from either axis (Fig. 4B,C, Supplementary Video). The ability to create such a representation provides a powerful, efficient and intuitive way of collecting H&E-like data from a patient and presenting it to a clinician, avoiding the randomness and imprecision of the sectioning process^24,25^, where only a few slices from an entire tissue block can be examined under a microscope.

**Figure 4:**
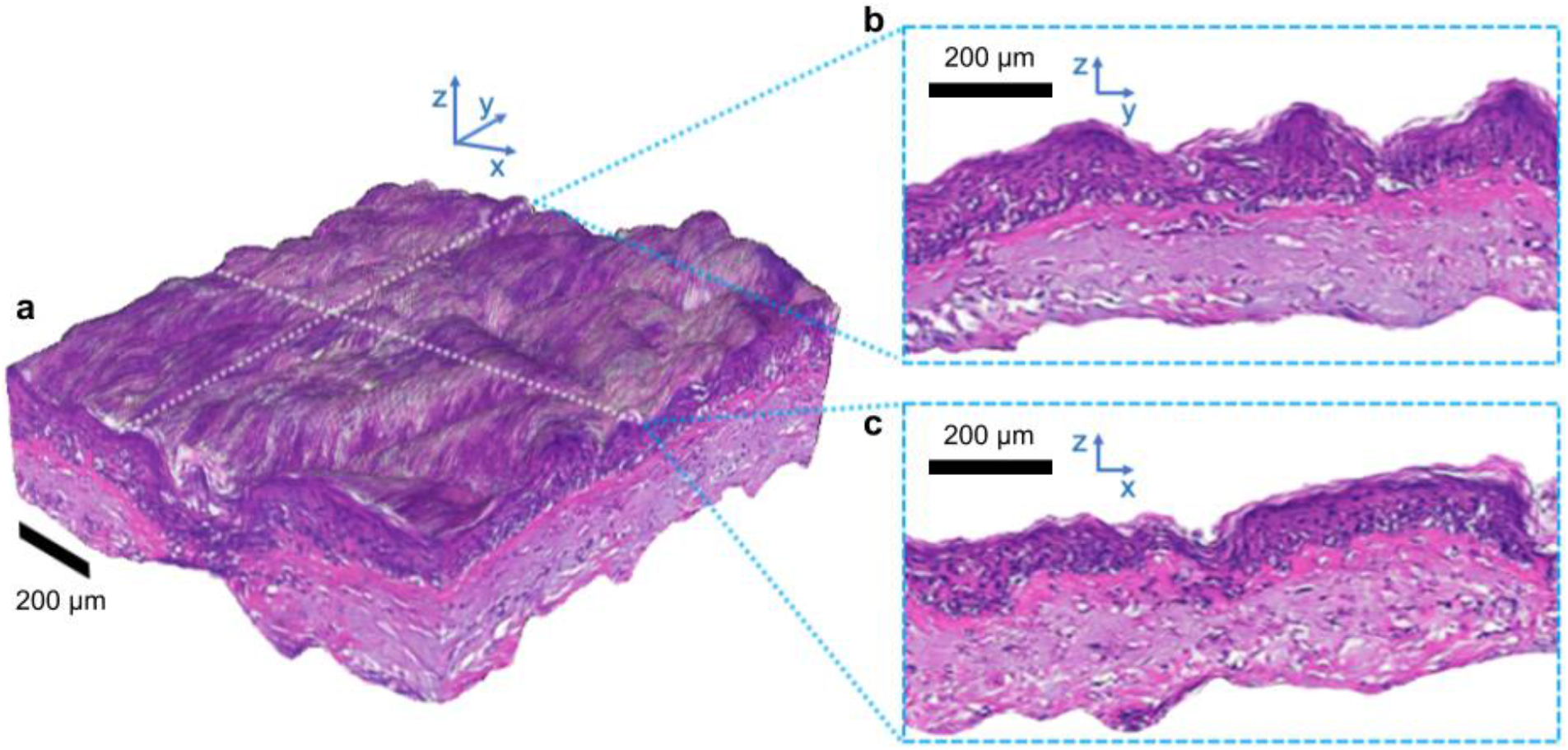
A 3D virtual H&E volume generated slice-by-slice from an OCT volume. The volume can be viewed in multiple ways, including isometric projection (a) and 2D cross-sectional views from within the volume (b, c), which can be chosen to slice across any plane.

## Discussion

Our work has demonstrated a proof-of-concept of the 3D virtual biopsy approach: the generation of highly realistic virtual H&E sections directly from OCT images. This result in turn relied on the feasibility of using our optical barcoding technique to create a large database of accurately aligned pairs of OCT and H&E histology images. These aligned image pairs were essential for generating high quality virtual histology images with our cGAN, and tests using an alternative cGAN that is trainable without aligned images generated output images of much lower quality (Fig. S12).

There are two primary directions for taking this work further, which we anticipate will open the door for a variety of basic research and clinical applications. First, optical barcoding can be used to obtain the precise H&E ground truth of OCT images as well as other types of optical image. Obtaining reliable ground truth enables multiple applications such as verifying the binding of contrast agents, better understanding how different structures may be distorted in histology, or for obtaining data needed to train other machine learning algorithms, such as ones designed to directly classify cancers or other abnormalities from an OCT image.

Second, our OCT2Hist process can be applied and further developed for a variety of basic and clinical research applications. Once it is properly trained, the image translator can be used to produce virtual H&E images without any need to optically barcode the OCT-imaged tissue. Thus it can be used non-invasively in vivo either to diagnose various superficial skin cancers or to confirm tumour margins, bypassing the invasive, time-consuming, and costly biopsy and histological processing procedure^1^. This is particularly important in cases where small tumours may have been completely eradicated by the biopsy and subsequent cascade of inflammation: There is no non-invasive modality available to determine whether surgery is still indicated without making an incision that can lead to scarring on the patient’s skin, which is particularly undesirable on the face^26^.

We note several shortcomings of the present machine learning algorithm. The first is that because we had to run on freshly excited tissues, we used a relatively small dataset to train our model of 1,005 images from 71 patients. Modern cGANs are trained on an order of magnitude more data, which makes the model more robust and generalizable to not-previously-seen samples. We expect our model to generate even more robust epithelial structures, textures and stromal features as the training dataset increases. As is well documented for deep learning applications, we expect the quality and robustness of OCT2Hist’s results to scale with the amount of data collected^27,28^.

A second limitation is the relatively small amount of information present in OCT images due to resolution, signal quality and contrast mechanism. OCT image resolution sets a lower limit on the size of features that can be reliably generated by the neural network. In practice, speckle noise^1629,30^ and multiple scattering^31^ can further obfuscate the useful information within a voxel. For this reason, in this paper we focused on generating large features (>50 microns) that have sufficient reliable information in the OCT image. Although our algorithm generates fine histology features such as cell nuclei, we believe that they should be regarded as general texture features and not interpreted as the exact position of individual cells as current OCT signal quality does not allow us to capture this information. In our training and testing, we specifically addressed the issue of lack of OCT signal in certain regions of the image (due to optical extinction) by masking out low-signal regions in both OCT and H&E images. The optical extinction also limits the imaging depth, which we typically observed was up to 450 microns for reasonable signal-to-noise ratio for the machine learning algorithm. We believe that future work based on OCT systems with higher spatial resolution, such as commercial systems with the capability to reliably image single cells^32–34^, will yield virtual H&E images of significantly higher quality. Most importantly, the principles developed in this work are easily translatable to such higher resolution OCT systems.

We believe that our OCT2Hist model can pave the way for a true virtual biopsy that is able to generate intuitive H&E-like images, supporting clinical decision-making in dermatology by identifying tumour margins and confirming tumour diagnosis. We further believe our optical barcoding method can shed light on the relationship between structures seen in OCT, confocal microscopy and other optical methods and a histopathological section of the exact same area.

An immediate next step will be to demonstrate that the model can robustly generate H&E-like features of superficial basal-cell carcinoma (BCC), including cancer margins, from OCT images. This is a favourable test case, as BCC clusters are typically hundreds of microns in size^21,35^, putting them well within the range of our system. As a preliminary result, we added a few cancer samples to our training set and show feasibility for non-invasive BCC detection (Fig. S18).

## Methods

Our image-alignment and cGAN-training pipeline featured several steps: encasing the tissue in fluorescent gel, OCT imaging, writing the optical barcode by photobleaching, computationally extracting the reslice parameters from the optical barcode, user-performed fine alignment of the OCT-H&E image pairs, quality assurance, stain color normalization, removal of low SNR regions from image, and finally machine learning training. Additional information is in the Supplementary Materials.

### Sample extraction

All participants provided verbal consent to participate in this study under IRB-24307. We received fresh tissue samples daily from Mohs surgery operations. Samples were stored and transferred in keratinocyte media consisting of DMEM/F12 containing penicillin/streptomycin (0.1%); fungizone (40 μg/ml); B27 (without vitamin A), epidermal growth factor (20 ng/ml), and basic fibroblast growth factor (40 ng/ml; Sigma) to support cell viability and were processed within 4 hours after excision. Prior to processing, tissues samples were transected to create a smaller size of approximately 0.5 cm x 0.25 cm x 0.5 cm.

### Gel encasement

Each tissue block was encased in a fluorescent gel. The gel was produced by mixing 0.03 g of Knox gelatin with water and 50 µL of 50 µM Alexa 680-NHS Ester dye in a glass flask, and mixing on a tilt table for 10 min. We added 0.06 g of agar and an additional 0.045 g of gelatin to the mixture before bringing the mixture to a boil in the microwave (about 10 s). The mixture was cooled for 10 s and then poured over the tissue sample resting in a cassette. After about 5 min at room temperature, the liquid mixture completely solidified to a gel.

### OCT imaging and barcoding

Each encased sample was placed on an XYZ translation stage (Thorlabs MT3-Z8) for imaging and barcoding. All images were collected with a Thorlabs Ganymede OCT system (Thorlabs GAN220). The optical fiber transmitting light to the scanhead was replaced with a custom-made wavelength division multiplexer with two input ports (for the photobleaching laser and the OCT source) and one output port. The OCT system was outfitted with a 10x, long-working-distance objective lens (Olympus UMPLFLN10XW) with an effective numerical aperture of 0.2, which was used with silicone oil immersion (*n*=1.4, which approximately matches the refractive index of skin tissue).

We collected 48 volume scans of 1 x 1 mm, where the optical focus was translated by 10 µm between each scan by a z-axis translation of the sample. All volumes were then computationally stitched together to yield an OCT volume with isotropic lateral resolution. After imaging was completed, a 650 nm laser diode (LP660-SF50) was activated (about 3.3 mW at output) to photobleach the optical barcode with 2 passes per line, at a line rate of 1 mm/15 s. Afterwards, an overview scan (Supplementary Figure S4) of total size 7×8 mm, consisting of 1×1 mm tiles scans was taken. The overview scan was used to determine a specific distance to cut into the tissue block in order to reach the location of the optical barcode.

### H&E preparation

The gel-encased, bar-coded tissue was then submerged in formalin solution for 24 h, and then transferred to 30% ethanol solution for about 2 h. The sample then underwent histological processing (protocol is shown in Figure S5), and the two-iteration cutting procedure (described in the subsection “Histological Processing & Tissue Sectioning” of the Supplementary Materials) designed to capture tissue sections in the most optimal location relative to the optical barcode. From each tissue sample we ultimately received 15 unstained tissue sections, which were imaged in a Leica SP5 microscope with a 633 nm emission filter to capture the fluorescent optical barcode on each section. The sections were then H&E stained and white-light images scanned at 20x magnification in an Aperio Digital Pathology Slide Scanner.

### Alignment

The H&E sections were aligned to resliced OCT images using in-house code that allows the user to select the photobleached lines from each fluorescent slide image. The alignment parameters computed directly from the algorithm constitute the “stack alignment”. We then performed a manual “fine alignment” step, where the resliced OCT image was fine-tuned to match the H&E image by adjusting the translation in 3 axes and in-plane rotation.

Images were filtered based on H&E image quality, OCT image quality, and alignment quality. We cropped OCT-H&E image pairs to 1024×512 pixels at 10x magnification (1 µm/pixel). We masked out areas of the image where the OCT signal did not have a sufficient signal to noise ratio to prevent the neural network from hallucinating tissue features where there is low signal in the OCT image. The results in this work were obtained by training the pix2pix model on the OCT-H&E image crops, which are automatically resized to 256×256 at runtime within the pix2pix preprocessing code. The model also outputs test results at 256×256, which are resized within an external code base to 1024×512.

The pix2pix model was modified to use the resnet9 generator, with 50% dropout, and with additional training-time data augmentation of random image translation. Use of the resnet9 generator (11.4 million parameters), compared to the baseline U-net network (54.4 million parameters), was found to improve generator mode collapse issues. The random translations, which consisted of random translations of up to 50% of the image’s length on each axis, were found to improve the quality of generated image features, such as hair follicles and epithelial structures.

## Data Availability

The data that support the findings of this study are available from the corresponding author upon reasonable request

## Acknowledgements

This work was funded in part by grants from the United States Air Force (FA9550-15-1-0007), the National Institutes of Health (NIH DP50D012179, K23CA211793), the National Science Foundation (NSF 1438340), the Damon Runyon Cancer Research Foundation (DFS# 06-13), Claire Giannini Fund, the Susan G. Komen Breast Cancer Foundation (SAB15-00003), the Mary Kay Foundation (017-14), the Skippy Frank Foundation, the Donald E. and Delia B. Baxter Foundation, a seed grant from the Center for Cancer Nanotechnology Excellence and Translation (CCNE-T; NIH-NCI U54CA151459), and the Stanford Bio-X Interdisciplinary Initiative Seed Grant (IIP6-43). Y.W. is grateful for a Stanford Bowes Bio-X Graduate Fellowship, Stanford Biophysics Program training grant (T32GM-08294). I.T. is grateful for NSF graduate fellowship. A.D.Z. is a Chan Zuckerberg Biohub investigator and a Pew-Stewart Scholar for Cancer Research supported by The Pew Charitable Trusts and The Alexander and Margaret Stewart Trust. K.Y.S. is the D. G. “MitcH&#x201D; Mitchell Clinical Investigator supported by the Damon Runyon Cancer Research Foundation (CI-104-19).

Additionally we would like to thank Anh Ngoc Diep, Murray Resnick, Bhargavi Garimella, Bryan Ronain Smith, Elliot SoRelle, Kent Lee, Lily Nguyen, Madeline Rose Hays, Max Prigozhin, Peng Si, Tommy Winetraub, Vivian Hua, Pauline Chu, Ziv Lautman, Jim Strommer and Graham P Collins.

## Author Information

### Author Contributions

Y.W. conceived of the presented idea. Y.W. developed the theory behind the alignment algorithm. Y.W., E.Y. and I.T. performed the computation of the alignment algorithm. E.Y. and Y.W. performed numerical simulations for the alignment. E.Y., I.T. and Y.W. developed the machine learning theory and performed the computation. Software supporting this project was developed by Y.W., E.Y., I.T. and E.B.. I.T., E.Y., Y.W., C.Y. and E.B. developed and performed instruments calibration. Y.W., E.Y., I.T. C.Y., S.S. and M.H. developed the gel embedding protocol used in the alignment step. S.A., W.C, H.D. collected samples from patients and coordinated secure transportation to our imaging facility. C.Y., S.S., M.M, J.Y., M.H. imaged and processed samples to generate the dataset used in this paper.

A.D.Z., K.Y., S.A., S.C. and K.E.R. contributed to the overall design and direction of the research.

The manuscript was written through contributions of all authors. All authors have given approval to the final version of the manuscript.

## Supporting Information

Supplementary Information is available for this paper.

Additional description of the materials, methods, and results; Figures S1–S18 (PDF). Movie S1 showing a 3D rendering of one of virtual histology volumes (MP4).

