## Supplemental Materials for "OCT2Hist: Non-Invasive Virtual Biopsy Using Optical Coherence Tomography"

### Supplementary Material

Our supplementary methods section is organized as follows:

1. Alignment Method
    a. Optical Design
    b. Photobleach Pattern
    c. Slide Alignment
2. Protocol
    a. Sample Preparation
    b. Scanning
    c. Histological Processing & Tissue Sectioning
    d. Fine Alignment
    e. Machine Learning Preprocessing
    f. Quality Control
3. Alignment Accuracy
4. Machine Learning
    a. How Lack of Alignment Impacts Results
    b. Dataset
    c. Additional Samples From Test Set
    d. 3D Volume
    e. Basal-Cell Carcinoma Preliminary Results

### Alignment Method

#### Optical Design

The imaging system used in this work consists of several components: a Thorlabs Ganymede system with a spectrometer imaging from 800 - 1000 nanometers. A sample translation stage system consisting of 3x Kinesis K-Cube Brushed DC Servo Motor Controllers and an MT3-Z8 3-axis stage system.  A custom Wavelength Division Multiplexer from FontSplitter which has 3 ports, two input ports for red (650 nm) and NIR (800-100 nm), and a combined output port.  A laser diode Thorlabs LP660-SF50 and compact laser diode/temperature controller Thorlabs CLD 1011LP.

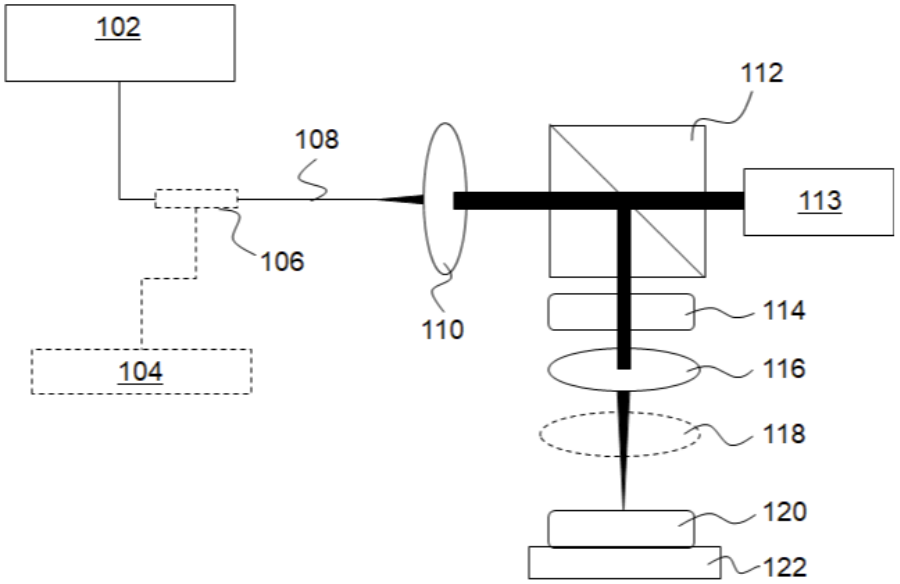


**Figure S1** Optical schematic of the OCT system.

We further describe the optical system in the following.  **Figure S1** shows an optical schematic of the OCT system used in this work.  Here 102 is a light source and spectrometer, 104 is an optional second light source and 106 is an optional fiber coupler for combining the outputs of sources 102 and 104. Fiber 108 carries light from source 102 to the rest of the system.  Collimator 110 collimates light emitted from fiber 108. For simplicity, the reference arm needed for an OCT system is shown schematically as block 113. Beam splitter 112 splits light from collimator 110 so that some light goes to reference path 113 and some light goes to sample 120. Sample 120 is mounted on translation stage 122. Between beam splitter 112 and sample 120 is an optical train including a galvanometer 114 (for scanning the beam), an objective 116 and (optionally) a removable spectral filter 118. In operation, light from sample 120 and light from reference arm 113 is combined by beam splitter 112 to interfere in a detector included in source 102. Galvanometer 114 provides 2D beam scanning, and it can be a combination of two orthogonal 1D scanners, or a single device that provides 2D scanning.

It is important to do the photobleaching of the dye at a different wavelength than the wavelength used for imaging. If the two wavelengths are the same, the OCT imaging undesirably creates its own photobleached pattern. However, it is convenient, and significantly more accurate, to use the OCT system to write the alignment pattern, as opposed to using a separate optical system to write the pattern. Thus it is preferred for the OCT system to be provided with a dual wavelength capability. There are two ways to do this.

The first approach is based on the use of spectral filter 118 in the beam path between source 102 and sample 120. If the OCT source is a super luminescent diode or a swept source, then we can utilize the system as following: we apply a low pass, high pass or notch filter 118 (depending on the light source wavelength relative to the dye absorption band(s)). The function of the filter is to ensure the light used for the OCT scan does not bleach the dye. Although **Figure S1** shows filter 118 disposed between objective 116 and sample 120, filter 118 can be disposed anywhere between source 102 and sample 120. For example, after objective 116, just before objective 116, before 2-D galvanometer 114, or before, after or within fiber 108. This 2D galvanometer can be provided by two 1-D galvanometers (our approach in this example) or with a single device providing both scanning directions. We chose to locate filter 118 after objective 116 because it requires minimal modification of an off the shelf OCT system. While we scan, the dye is not bleached by the laser because the wavelengths that the dye is sensitive to are shielded by the filter. After the scan we remove the filter, increase the OCT laser power and use the same system to photobleach. The second time around the part of the OCT spectrum that the dye is sensitive to reacts and photobleaches the dye.

The second approach is applicable in cases where the OCT system utilizes light that the dye is not sensitive to. Here we need to couple another light source (i.e., optional second light source 104) and turn it on when we want to photobleach (here removable filter 118 is not used). Here optional coupler 106 is used as wavelength combiner in the forward direction (left to right) and as a wavelength splitter in the reverse direction (right to left). This ensures that high optical intensity as needed for photobleaching does not enter the OCT detector system within block 102 (which could be damaging to the detector). It also ensures that the OCT signal light is routed to block 102 for detection.

The first approach is better if there is an overlap in wavelengths between dye absorption bands and OCT emission wavelengths, and the second approach is better if there is no such overlap. The second approach has better vertical resolution than the first approach because we don't block any of the OCT light. Both methods guarantee that photobleaching is done in the same coordinate system as OCT scanning and both methods are better than using a completely separate way to photobleach. Practical systems will include the second light source 104 or the removable filter 118, but do not need to have both.

Another practical issue is the translation stage. Because we use a high NA optical system we can't have the entire sample in focus at the same time. In principle there are two ways to deal with this: either by moving the sample, or by moving the OCT scanning head, another alternative is to design an optic element that will shape the beam as a needle such that the entire sample is in focus (for example Bessel beam), or that have multiple focus positions in the sample. We choose moving the sample.

This way turns out to be easier for correcting for path length aberrations in the index of refraction mismatch between the tissue and the medium around it thus increasing system stability and resolution. Here the measure for stability is displacement in x,y,z as a result of the act of imaging.

Thus translation stage 122 is preferably a 3D translation stage that can travel in three orthogonal directions.  It can travel vertically to bring the sample into focus, but can also travel in two transverse directions to bring different parts of the tissue into the field of view of objective 116.

#### Photobleaching Laser optical fiber discussion

In addition, an external laser of a single wavelength was connected to the OCT system through a designated optical fiber (custom WDM, 3.3 mW at exit) in order to photobleach the sample. We customized the fiber in a way that allows the transmission of the OCT signal and the single wavelength laser with minimal power loss.

Another important aspect of this example is the scanning parameters. Using the system described above, we tested different lateral pixel sizes (1 µm and 2 µm) and chose 1 micron pixel size, which is smaller than the spot size of the OCT beam) that enables enough information on any position in the sample.

### Photobleach Pattern

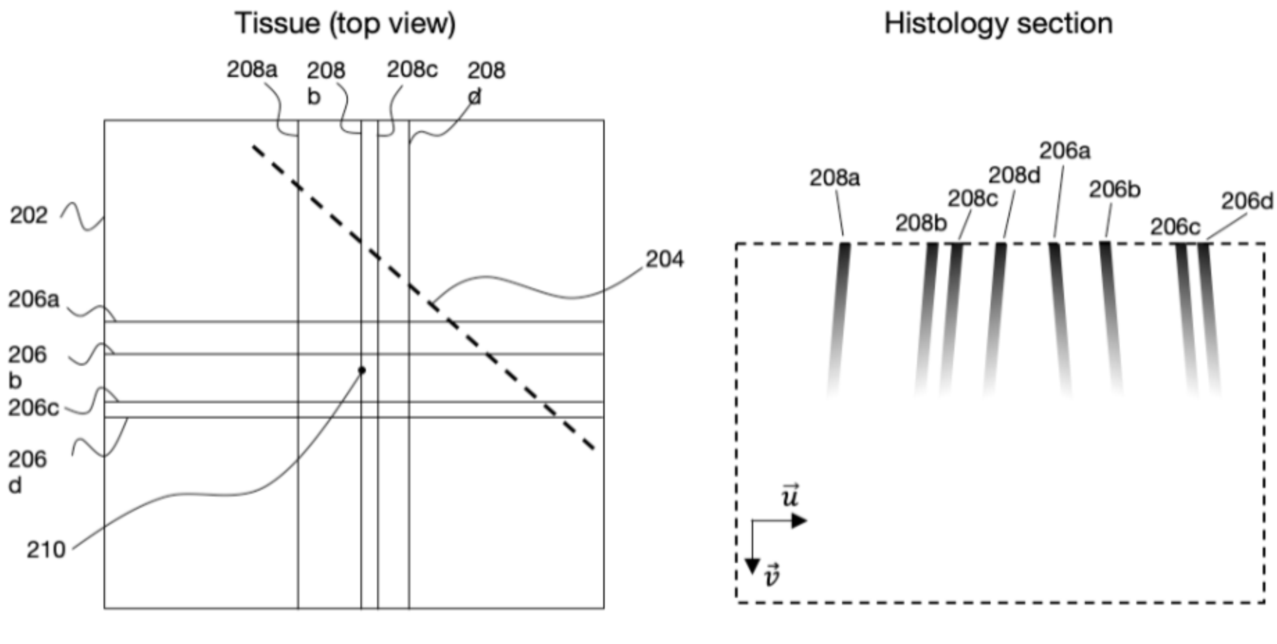


FIG. S2A FIG. S2B

**Figure S2** Several examples of the optical barcode pattern photobleached onto the surface the fluorescent gel which encases the tissue. (a) A view from above (looking down on the sample) of an optical barcode with 4 lines in vertical direction, 206a, 206b, 206c, 206d, and 4 lines in the horizontal direction, 208a, 208b, 208c, 208d. A histological section is taken along the dotted line. (b) The side view (u-v plane) of the histological section, showing the photobleaching in depth for each of the 8 lines in the barcode.

Figure S2 shows a top view of an exemplary 8-line pattern (4 of each kind). This is the kind of pattern that was used in our experimental work. Here 202 is the OCT field of view, 210 is the origin (i.e., OCT scan center), 204 is the histology cut, 206a, 206b, 206c, 206d are horizontal pattern lines as described below and 208a, 208b, 208c, 208d are vertical pattern lines as described below. As indicated above, line spacings are preferably chosen such that the identity of each line is encoded by the spacing of that line to the surrounding lines (the distance ratio between the surrounding lines is unique). In our current implementation we use 4 horizontal and 4 vertical lines. Horizontal lines are positioned at -3*b, -2*b, 1*b, 3*b and vertical ones are in positions -4*b, 0, 1*b, 3*b. Here the origin (0,0) is the OCT scan center, and b is the base unit of measure and depends on the field of view being encoded (we choose 100 microns but 50 microns up to 200 microns will also work). Note that the resulting distance ratios are preserved independently of the angle the histology cut makes with respect to the lines. Uniform shrinkage of the sample during histology preparation also has no effect on these distance ratios.

The preceding is an example where the predetermined pattern is formed by writing a top surface of the 3D sample with a pattern of intersecting lines, whereby the predetermined pattern is a corresponding pattern of intersecting planes. The pattern of intersecting lines preferably includes a first set of lines perpendicular to a second set of lines. The first set of lines preferably has a first spacing pattern between the lines that permits identification of a line from its distance ratios to adjacent lines in the 2D image (as in the example of **Figure S2c**). The second set of lines preferably also has a second spacing pattern between the lines that permits identification of a line from its distance ratios to adjacent lines in the 2D image (as in the example of **Figure S2c**). E.g., in the example of **Figure S2c,** a line in the histology image having a distance ratio of 1:2 with respect to its neighbors can be identified as coming from line 208c of the pattern of **Figure S2c.** A line in the histology image having a distance ratio of 2:3 with respect to its neighbors can be identified as coming from line 206b of the pattern of **Figure S2c**. Edge lines of the patterns (i.e., 206a, 206d, 208a, 208d) can be identified based on their positions relative to lines that can be identified via distance ratios (i.e., 206b, 206c, 208b, 206c).

The position of the alignment markers should be set in a way that the difference in distance between consecutive lines d1,d2,d3,... will be unique. Where d1 is the difference between line 1 position and line 2 positions, etc. For simplicity, we can assume d1,d2,... are integer numbers and there is some base unit b (in microns) that multiplies d1,d2,... to get the exact separation. In order to select b we need to balance between smallest separation between two lines - we don't want it to be comparable with the photobleach laser spot size for example, because then we can't tell lines from one another. We also don't want b to be too big because the lines will be far apart and outside the field of view of the laser, so we cannot photobleach all of them. For our setup we choose b=100 microns, but values from 50 to 200 also work well.

There are a few ways to select d1,d2,... for example by setting each of them to be a different prime number, by doing so, we can uniquely know which line we are looking at by comparing the distance to its neighbors. Using a different prime number for each distance is not efficient in the sense that prime numbers grow relatively quickly which means difference between consecutive lines will become larger and larger until lines are out of the field of view.

A more efficient way to encode, is to allow for the same integers to repeat but to prevent the same division of number to repeat di/d(i+1) only once. This problem can be written as a graph problem in computer science by creating a graph where every vertex (1,2,3,...,V) represents a difference di and two vertices are connected if their greatest common denominator is 1. For example there will be an arc between vertex 2 and vertex 5 but not between 2 and 4. We then need to find a connected path that has n steps (n+2 being the number of lines required, n+1 is the number of differences, hence n is the number of arcs in the graph). This turns into an optimization problem: find an n step path with the penalty being the sum of all the vertex passed, every edge of the graph can be gone through once. Minimizing this penalty will yield the following differences. Each has a unique encoding of di/d(i+1).

3 lines: H lines:1-2. V lines:1-4.

4 lines: H lines: 2-1-4. V lines: 2-3-1

4 lines second example: H Lines: 1-2-3, V lines:4-3-1. The 4-lines second example is slightly better as more of the sample is covered by lines.

5 lines: H lines:1-2-5-1, V lines 1-3-4-1

#### Slide Alignment

The histology plane estimation problem is mathematically formulated by parametrizing the histology plane and then solving for the parameters of that plane. Every point on the histology plane satisfies the following equation:

$$\vec{p} = u\vec{u} + v\vec{v} + \vec{h} \qquad (1)$$

where $u_x,u_y,u_z,v_x,v_y,v_z,h_x,h_y,h_z$ (i.e., the components of , , and ) are the parameters of the plane to be estimated. The vectors u and v describe the plane, such that any point on the plane can be written as a linear combination of vectors u and v. The vector h points from the origin of the coordinate system to the left hand corner of the u-v plane.

We can also not approximate the histology as a single plane, but as a curved manifold or a collection of multiple planes. One will do so by dividing up the histology section into multiple parts, and use lines present in each part to locally approximate a plane. Because we only need 2 horizontal lines and 2 vertical lines to compute a plane, and since we have 3 or more lines of each kind, we can fit multiple planes say one plane for the left hand side of the histology and one plane for the right hand side and perform 3d interpolation between the different parts of the histology section. We saw that there isn't much difference between these planes so it is better to fit just one plane, as the error from that is very small.

We can identify specific points in the histology plane ($u=u_1,v=v_1$), ($u=u_2,v=v_2$) etc. that are both on the histology plane (by definition) and on the photobleach lines. We can also identify which of the photobleach lines ($L_1$, $L_2$, etc.) by using the photobleach position ratio as described in more detail below. In this example we assume ($u_1,v_1$) is on line $L_1$, ($u_2,v_2$) is on line $L_2$ etc. We can estimate some of the plane parameters by using least squares method or otherwise providing an optimal solution to this system of equations:

$$\begin{pmatrix} L_{1x} \\ L_{2x} \\ L_{3y} \\ L_{4y} \\ \cdots \end{pmatrix} = \begin{pmatrix} u_1 & 0 & v_1 & 0 & 1 & 0 \\ u_2 & 0 & v_2 & 0 & 1 & 0 \\ 0 & u_3 & 0 & v_3 & 0 & 1 \\ 0 & u_4 & 0 & v_4 & 0 & 1 \\ \cdots & \cdots & \cdots & \cdots & \cdots & \cdots \end{pmatrix} \begin{pmatrix} u_x \\ u_y \\ v_x \\ v_y \\ h_x \\ h_y \end{pmatrix} \quad (2)$$

This system of equations is formed by taking x and/or y components of Eq. 1 for each point used for the fitting, and noting that points on the photobleach lines have known x or y coordinates which appear on the left hand side of Eq. 2. Briefly, $L_{1x}$ etc. are known from the pattern on the top surface of the sample, and $u_1$, $v_1$ etc. are read off from point positions in the histology plane. Preferably, enough points are used to make the system of equations overdetermined.

This solves the estimation problem for 6 out of the 9 parameters. The remaining 3 are $u_z$,$v_z$ and $h_z$. The parameters $u_z$ and $v_z$ can be estimated by assuming no shearing and uniform shrinkage:

$$\vec{u} \cdot \vec{v} = 0 \quad (3)$$

and

$$\|\vec{u}\| = \|\vec{v}\| \quad (4)$$

We could alleviate these constraints by allowing some shrinkage and shearing, which converts Eqs. (3) and (4) to

$$\vec{u} \cdot \vec{v} = a \qquad (3b)$$ and

$$\|\vec{u}\| = \|\vec{v}\| + b \qquad (4b)$$

where a and b are free parameters of the problem, estimated for each alignment. However, we found that for most realistic cases their value is very low, so we can assume a=0 and b=0 thus simplifying the equation.

Finally, $h_z$ can be independently determined (e.g., by fitting the tissue surface between histology and the OCT). In order to determine $h_z$ position we used manual registration of tissue surface with the OCT surface. We considered a few options to do so: 1) matching tissue surface seen in OCT and seeing in histology, 2) matching gel surface between two modalities, 3) matching features inside the gel, e.g. natural occurring clumps or adding beads to the gel to provide as landmark to allow us to adjust $h_z$ until match is formed, 4) using the Gaussian shape of the beam as it is photobleached into the gel to estimate what depth was the focal point at the time of bleaching. We choose option #1 for simplicity, it also doesn’t require one to find any specific features at the gel and is less sensitive to shearing or morphing of the gel itself.

We also use the marking of tissue interface as a proxy for alignment quality, by measuring the amount of overlap and distance between tissue interface as seen in OCT and histology - we can estimate how far the received alignment is from the actual alignment.

Since we request multiple sections and slides from histology, we make the reasonable assumption that all sections coming back from the same batch (or as we call it, iteration) 1) have the same spacing between them and 2) have the same plane normal. We can use multiple sections from the same iteration to further fine tune the plane parameters. We do so by averaging the normal vector of all the planes to get a stack normal vector, we also average the uniform shrinkage factor $|u_{section}|$ to get a stack uniform shrinkage factor $|u_{stack}|$.

Finally, we compute the plane distance for each section to OCT origin and fit a linear line. Our assumption is that on average, histology planes are equally spaced along the cutting direction.

After computing average normal, average scale factor |u| and fit a linear distance from origin, we can recompute plane alignment for each slide - this will be considered as the stack aligned planes. One can repeat the same method but extracting outlier planes such as planes that have unusual scale factor (sample usually shrinks by 5% to 15%, so sections that shrink further, or expand are considered suspicious), or do not fit with the linear plane distance fit (say more than 50um away from the linear fit), or have their normal have large angle compared to the stack normal (more than a few degrees) and recompute for higher accuracy.

Following the stack alignment step, we give the user the ability to fine tune the alignment using a simple graphic user interface. The user can change uniform scale factor, rotation angles and position of each slide individually by comparing the OCT image with the corresponding histology and finding features that appear in both such as tissue structure and curvature, hair follicles, cell clusters, gel clumps, etc.

### Protocol

#### Sample Preparation

**Figure 2** in the main text is an exemplary flow chart for providing paired OCT and histology data according to the OCT2HIST method. We received fresh tissue samples daily, in the afternoon, from Mohs surgery operations in the morning. The tissue samples are approximately 1 cm x 0.5 cm x 0.5 cm in width x height x depth and immersed in tissue media after surgical excision.

##### Encasing Sample in Fluorescent Gel

We cut the sample to an appropriate size for imaging of about 0.5 cm x 0.25 cm x 0.5 cm and encase the sample in a custom-designed fluorescent gel. The gel is produced by first mixing 0.03 g of Knox gelatin with water and 50 uL of 50 uM Alexa 680-NHS Ester dye in a glass flask, and mixing on a tilt table for 10 minutes. We add 0.06 g of agar and an additional 0.045 g of gelatin to the mixture before bringing the mixture to a boil in the microwave (about 10 seconds). The mixture is cooled for 10 seconds before being poured over the tissue sample resting in a cassette. After about 5 minutes at room temperature, the liquid mixture should completely solidify to a gel.

#### Scanning

##### Sample Orientation

The tissue sample sample is typically cut to a rectangular shape, where the long axis is approximately twice the length of the short axis. During imaging the sample is oriented such that the long axis is at 45 degrees with respect to the internal x-y-z coordinates of the OCT system. This guarantees that when the sample is sectioned along the long axis, the cuts intersect both horizontal and vertical lines of the optical barcode at about 45 degrees, as shown in **Figure S3**.

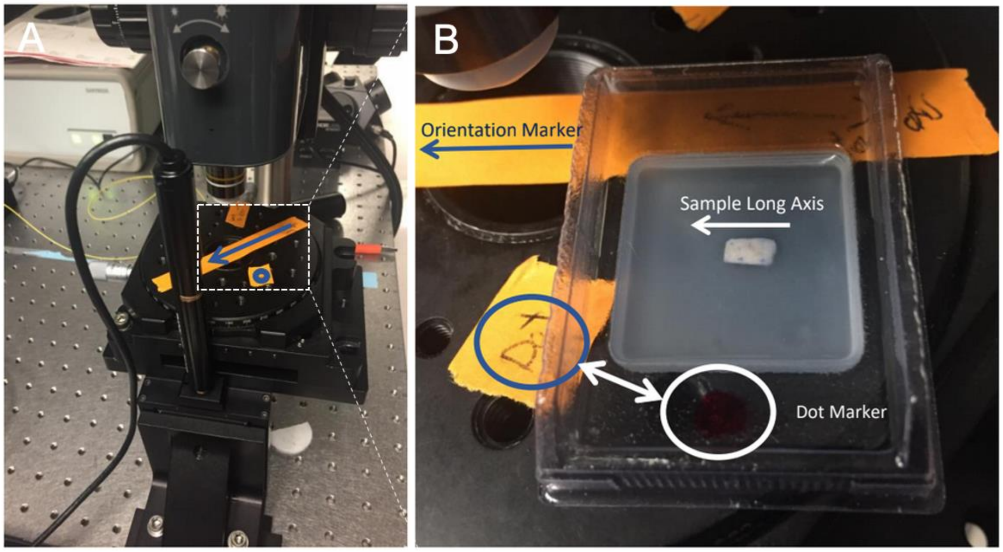


**Figure S3** (a) Picture of our OCT scanning head and imaging stage, which is mounted on a motorized 3-axis translation stage. The arrow marked on the orange tape shows the desired orientation of the sample long axis so that it is oriented at 45 degrees relative to the OCT coordinate system. (b) Shows a zoom-in of the stage with the tissue sample embed in gel and correctly oriented.

#### OCT z-stitching discussion

The high-quality stitched image requires the acquisition of multiple images in different focus depth locations. In order to do so, we can either move the sample or move the OCT scanning head. We have decided to move the sample rather than the OCT scanning head, in order to maintain the same physical focal plane Z location, and as a result to improve the stitched image quality and simplify the stitching algorithm, because the stitching algorithm performance is dependent on correct estimation of the focal plane position. In order to translate the sample, we have placed it on a moving stage (X,Y and Z translation) and optimized the Z stage steps, by minimizing the scanning time while sampling tight enough (the step size is smaller than the effective Rayleigh length), to preserve high lateral resolution. The step size used for OCT images in this work is 10 microns.

In addition, to prevent tissue degradation we start by scanning the tissue from top to bottom, and only after we photobleach the markers. The sample was also immersed in silica oil (rather than water) to better match the tissue index of refraction, and to prevent the tissue from drying. The scanning field of view was chosen in the flattest area of the middle of the tissue sample, to avoid the distortions that occur at the tissue sample's edges and causes unwanted deformations (due to shearing) of the photobleached lines. In addition, we started the acquisition 190 microns above the tissue surface and ended it 500 microns below it. This ensures that even if the tissue is slanted, we will still capture the surface while maintaining sufficient depth imaging.

In addition, the field of view was chosen to be 1 mm by 1 mm. We optimized this parameter by drawing 2 hashtags (total of 8 photobleached lines) in two separate locations (center to center distance of 500 microns). We then used all possible lines combinations (4 lines are needed to estimate a plane), to estimate all possible planes. We found that within the 1 mm by 1 mm range all plane estimations were similar, which indicates minimal deformation/bending of the gel in that region due to histology cut, resulting in more accurate plane estimation. Field of view was also one of the parameters we optimized for while selecting a lens.

#### Optical Path Correction

Because the galvos of our OCT system are not close to the focal position of the imaging lens, the optical path length varies as a function of scan x,y position, and the OCT image exhibits an apparent curvature even when imaging a flat surface. We digitally compensated for this effect by measuring a surface that is physically almost completely flat, the bottom of a glass petri dish. We inverted the OCT-measured curvature of this glass petri dish to normalize the optical path length across the image. We fitted a quadratic equation in x and y that approximated the measured curvature to less than 0.5 um error on average. Although the exact path length correction is a function of depth, we found the measured path length change to be almost constant in depth. Hence, we use a single correction for all depths.

#### Sample Overview

After imaging and photobleaching the tissue sample, we perform the sample overview, which is used to obtain the first iteration of tissue sections. The sample overview consists of tiled, low resolution OCT scans performed at two depths, all of which are stitched together to give the user an en-face "overview" of the entire sample, and the optical barcode's position within the sample. The overview consists of 8x7 OCT volumes with pixel size 50 um (laser spot size is the same as the regular volume, 50 um is the jump between pixels) and field of views 1 mm by 1 mm each acquired at 3 depths of equal spacing between 190 um above the gel tissue interface and 1 mm under the gel tissue interface. All tiled scans are stitched together to form an 8 mm x 7 mm volume (**Figure S4**) and presented to the user, who will then choose a side of the sample to begin cutting from.

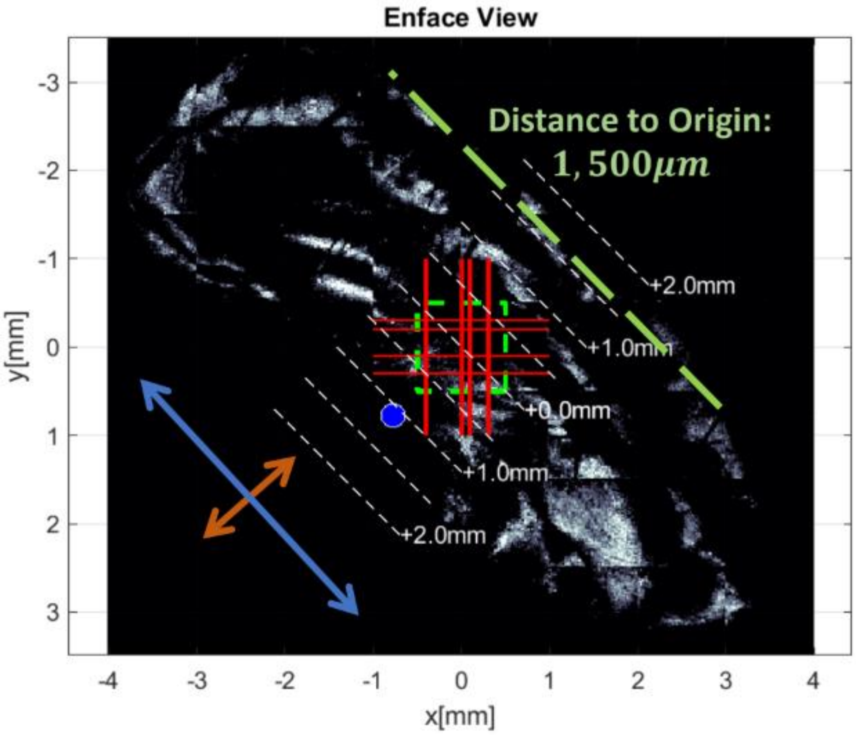

**Figure S4** Overview scan taken after OCT imaging and photobleaching. The entire tissue sample is visible with the optical barcode position overlaid. The distance to the optimal zone for cutting, which is between -0.5 mm and +0.5 mm, can be measured from the edge of the tissue.

### Histological Processing & Tissue Sectioning

The scanned and photobleached tissue is submerged in formalin solution for 24 hours, and then transferred to 30% ethanol solution for about 2 hours. The histologist subjects the sample to multiple steps of tissue processing as shown in the image below with the result that the tissue is encased in paraffin wax.

It is difficult for the histology sections to be cut at the desired position with sufficient accuracy to provide training data as described above for machine learning to provide synthetic histology. This difficulty is addressed by a first iteration of the alignment method that provides information on the location of a first histology plane in the OCT image. This provides a reference such that the second histology series of cuts can be specified relative to the first histology iteration to ensure the second histology iteration goes through a useful part of the sample for training data. Thus the instructions for the second histology iteration can be something like "cut to specified depth, relative to the first iteration.

After cutting the first iteration of tissue sections, the paraffin wax is melted off before fluorescent imaging. After cutting the second iteration of tissue sections, the paraffin wax is melted, fluorescent imaging at 633 nm is performed in the Leica SP5 confocal microscope at 10x magnification, sections are then H&E stained, and then scanned at 63x resolution.

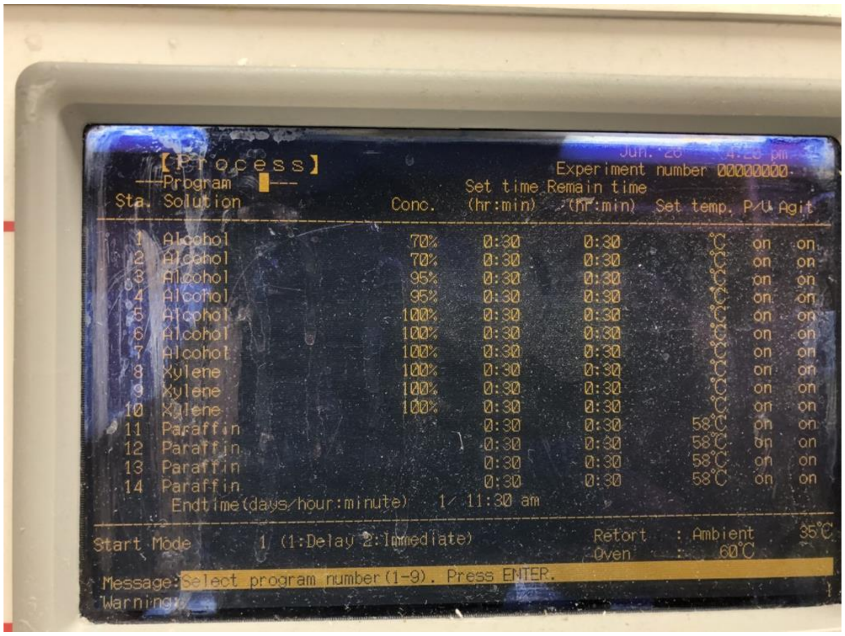


**Figure S5** Picture showing the tissue processing and paraffin embedding solutions and schedule.

#### First Iteration

The user typically selects the cutting side to be that in which the optimal zone of the optical barcode is closer to the edge of the tissue sample, which allows the histologist to shave through less tissue to reach the optimal zone. The histologist is instructed to take 6 tissue sections upon cutting to a full-face of the sample, starting from the cutting side previously specified. The 6 tissue sections are then fluorescence imaged, capturing their respective barcode patterns. The sections are aligned to determine their location within the sample, and to determine how far they are from the optimal zone of the barcode.

#### Second Iteration

Based on the results of the first iteration, the histologist is instructed to cut further into the sample in order to precisely reach the optimal zone. He/she will then take 15 tissue sections of 5 micron thickness spaced 10 microns apart.  The second iteration sections are first fluorescence imaged before H&E staining.

#### Fine Alignment

Tissue sections from the second iteration are aligned by stack alignment.  After the stack alignment is done, the user is given the option to manually fine tune the alignment by moving along the OCT volume perpendicular to the aligned plane, and within plane. In this step, the  user will try to find the same features in histology and OCT and match tissue structure as well as structures inside the tissue and the gel.  We have developed a MATLAB-based user interface in which the user can view the merged OCT-H&E images with transparency, and tune the x-y-z translation of the OCT reslice, and the in-plane rotation.  The first step typically consists of aligning the surfaces of H&E and OCT image by tuning the in-plane translation and rotation, and then identifying idiosyncratic tissue features that can be matched in OCT-H&E to tune the out-of-plane translation.  Examples of this are found in **Figures S12** and **S13**, where we use two specific features, Rete pegs and a hair follicle to achieve fine alignment.

best aligned in panel b), and a shift of 25 microns in either direction reduces the quality of alignment.

There are additional methods to perform fine alignment for example brute force through many planes, maximizing the correlation between OCT and histology images, or align rotation in u-v plane then go through different positions perpendicular to the plane, feature matching between features on the tissue in OCT and histology, using beads or clumps in the gel to try and match as many. We found that manually fine-aligning the images, by adjusting the translation, rotation, and section y-position, yields the most accurate alignment results, in terms of both cross plane and in plane errors. We found that most samples have cross plane error of 20-25um from the stack alignment to the fine alignment, giving a bound on the amount of cross plane error.

An alternative method, as discussed above, is to use fluorescent beads which can be embedded within the fluorescent gel (at a different emission peak) encasing the sample. Due to their reflectance and fluorescence properties, the beads can be detected in both the OCT system and the fluorescent microscope, and can serve as a mutual feature in both imaging modalities. After the alignment algorithm estimated the plane and pulled the corresponding slice from the OCT volume, the particles can be used in order to:

1) Fine tune the estimated plane by maximizing the number of paired particles that can be seen in both images (fluorescent and OCT) and/or

2) Quantify the alignment performance, by estimating the transformation between the algorithm estimated plane and the fine-tuned plane.

Both uses are particle size dependent. Smaller particle size will result in better fine tuning and better alignment performance quantification. There are several metrics that can be used to determine how good our alignment is. Let us assume our best fit for the histology plane is H1, and the ‘real’ plane is H0, we can use the following metrics to determine how 'far' H1 is from H0.

1) cross plane alignment (we use this one most commonly): by determining how much H1 needs to move along the normal to H1 to match the average position of H0. We use this method as it describes an error which is not trivially visible when comparing OCT and histology images

2) Angular error - computing the angle between H1 normal and H0 normal. This angle is usually very small and so we rarely use it

3) Average point translation. We can map each point in H1 to a point in H0 and ask what is the average, median or max distance between these points.

#### Machine Learning Preprocessing

We apply a few additional processing steps to the paired images before the machine learning training. The first is digitally extracting a 5 um to 20um averaged slice of the OCT image for each section to match the physical 5 um H&E slices and cross plane uncertainty in the alignment. We only consider images within about +- 550 um of the center of the OCT scan as they have significant overlap between OCT and histology, in order to ensure each image is sufficiently large. We then perform several steps of masking on the OCT image. We crop the image to 100 um above the interface of the tissue in the OCT image, and 500 um below due to low SNR of OCT image which interferes with the learning process. One can choose other bounds depending on the details of the OCT system being used: depth of penetration namely. Additional masking is performed to crop out signal poor regions of both images, as these regions do not contain useful training data. We also resample both images to have the same scale of 1 microns, but other resolutions are acceptable. In choosing the resolution one should consider OCT imaging resolution and beam size, histology imaging resolution as well as the size of the features they are trying to reconstruct in the synthetic image. We find all OCT images can be vertically fit within a frame of 672 pixels, and horizontally fit within a frame of 2048 pixels. Hence, for each image we take two 1024x512 crops aligned to either horizontal end of the original image.

We perform a final stain normalization step on the H&E images. The individual H&E images exhibit a wide range of color spaces, even though each image technically contains only two stain colors for haematoxylin and eosin. There are a variety of experimental reasons why the staining colors can vary from sample to sample including stain concentration, staining method, and time. The end result is that some images have much more contrast, or deeper colors compared to others. This can confuse the machine learning algorithm.

We seek to overcome this problem  with stain normalization .  We use a SVD decomposition[1] of the pixels in each H&E image to find the 2 vectors capturing the largest amount of variance, which should correspond to the colors of haematoxylin and eosin.  With these vectors, we can extract the concentrations of haematoxylin and eosin at each pixel, and recalibrate the image with a new pair of H&E vectors.

#### Quality Control

We implement numerous steps of quality control to ensure that our training dataset of paired OCT and Histology images is of high quality. Much of this quality assurance data is collected through a digital survey, from the user performing the fine alignment.  The first check is of whether or not both the OCT and Histology images are of high quality.  For the OCT image, this means there is no significant shadowing effect, or a lack of contrast in the tissue, and the epithelium is distinguishable.  For the histology image, this means there is not significant tearing, damage to the tissue, or staining artifacts. We then rate the alignment quality between the H&E and OCT images, the following scale: “3 - Perfect Alignment”, “2 - Good, With Flaws”, “1 - Bad”, and a 0.5 point increment.  We only use images with a score of “2” or higher for constructing the machine learning database. We also gather additional statistics on the error of the y-axis (cross plane translation) fine-alignment by asking the user to rate how far (in um’s), he or she can shift the OCT image before it becomes obvious that alignment is wrong by looking at matching of features between the two images.  This allows us to gather statistics on how robust and useful the fine-alignment process is.

### Alignment Accuracy

We attempted to verify the accuracy of our alignment method by various methods.  The first is by aligning fluorescent beads of 25 micron diameter encased in a gelatin gel.  The beads are visible in OCT, fluorescence confocal microscopy, and in histological sections after the full histological processing.  We can create a fluorescent gel, embed it with fluorescent beads of a different emission color and compare the OCT image with an aligned image of a different modality.  In **Figure S6**, we image the gel with beads in a confocal microscopy right after OCT imaging.  We align the fluorescence image to our OCT volume using an optical barcode.  We observe that the same beads, visualized in both OCT and confocal microscopy, align almost perfectly.

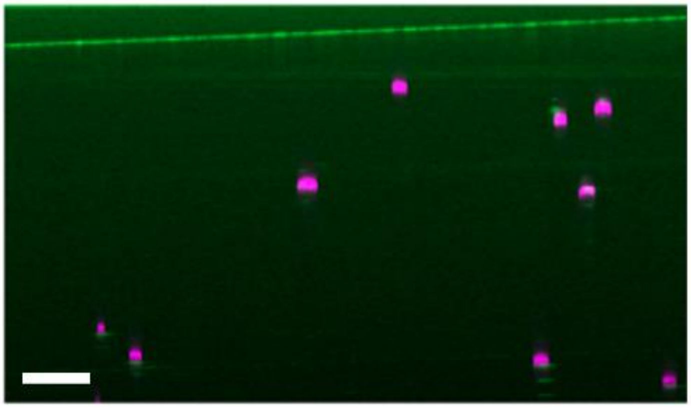

**Figure S6** Alignment precision verification using 25 micron PLGA beads embedded in gelatin gel. (a) OCT image of beads in green is aligned to a confocal microscopy image of beads in pink.

We can take this process step further, and observe the beads in a histological section, after undergoing the entire chemical processing pipeline that is typical for tissues (**Figure S7**). The resulting alignment is also very precise, with the same group of beads clearly observed in both OCT and histology section. We have successfully reproduced this bead experiment multiple times and thus, this is one method through which we have concluded that our alignment algorithm is capable of alignment precision down to 25 microns.

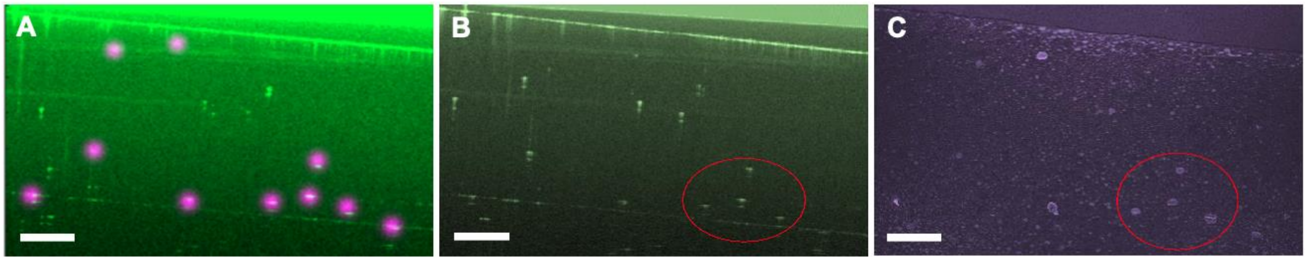


**Figure S7** Alignment precision verification using 25 micron PLGA beads embedded in gelatin gel. (a) OCT image of beads in green in aligned to histological image of beads highlighted in pink. (b) Histological image of beads in gel from (a). (c) OCT image of beads in gel from (a).

We can also observe the success of the optical barcoding technique by examining aligned OCT-H&E image pairs. **Figure S8** shows two image pairs of freshly excised human skin from Mohs surgery. The remarkable similarity between the OCT and H&E images demonstrates the quality of the alignment. The epithelial strand cell features pointed to with arrows in **Figure S8a,b** are about 50 microns in size. There are also well aligned larger features in the gel ranging from 100 to a few hundred microns.

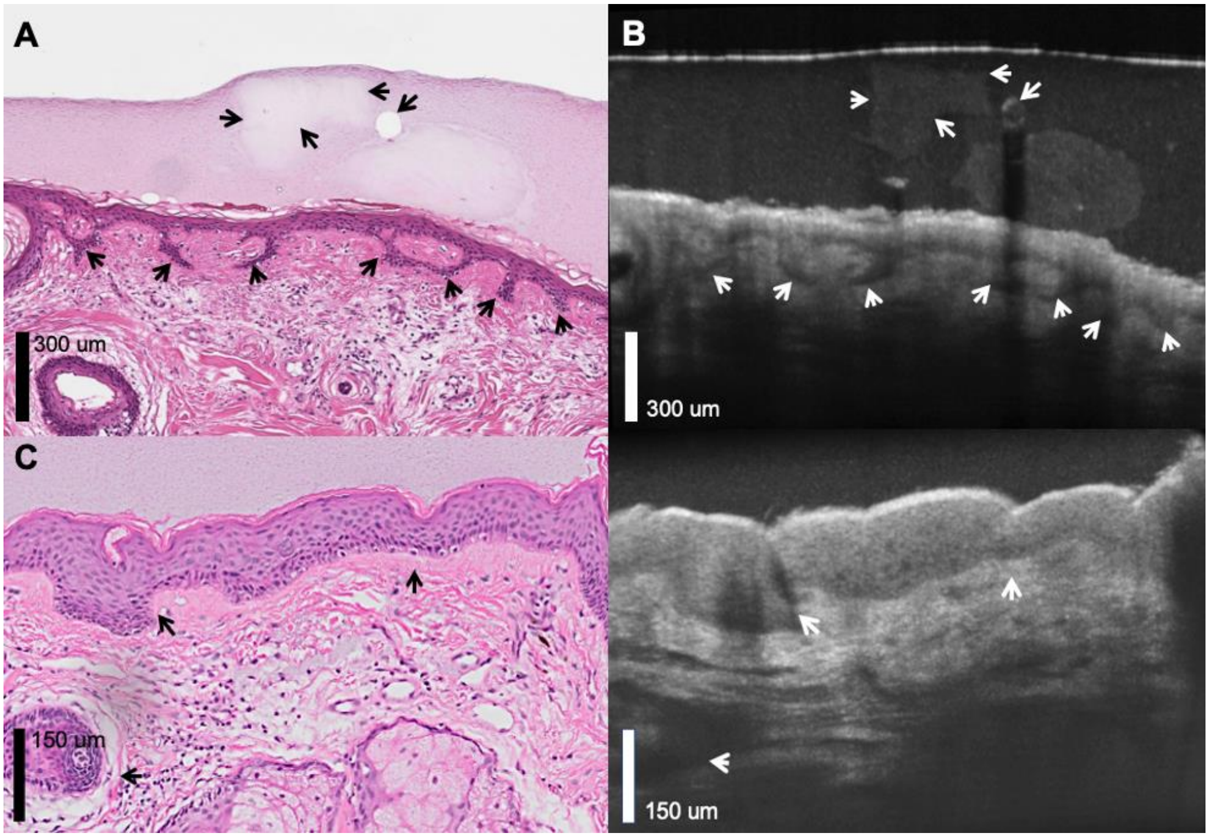


**Figure S8** Aligned OCT-H&E image pairs. (a,b) Aligned image of human skin from a male, age 66, scalp, obtained by Mohs surgery. The arrows point to various aligned features, including a clump in the gel, and strands of epithelial cells. (c,d) Aligned image of human skin from a male age 74, forehead, obtained by Mohs surgery. Arrows point to similarities in the epithelium, as well as a clump of cells in the dermis.

The last way in which we verify the precision of the optical barcoding technique is by comparing the stack alignment (the plane extracted from the alignment algorithm) with the fine alignment (the plane that is fined tuned by the user using various tissue and gel landmarks). In **Figure S9** we compile a list of tissue sections plotted against the out-of-plane distance between stack and fine alignment for each. The important numbers for assessing alignment accuracy are the error bars which indicate the user reported uncertainty in the fine alignment - literally it is out-of-plane distance in either direction the user can translate the OCT image before observing a discrepancy with the H&E image.

**Figures S10** and **S11** also illustrate that our fine alignment achieves approximately 25 microns in out-of-plane accuracy by comparing how tissue landmarks change if the OCT stack is shifted by 25 microns in either direction.

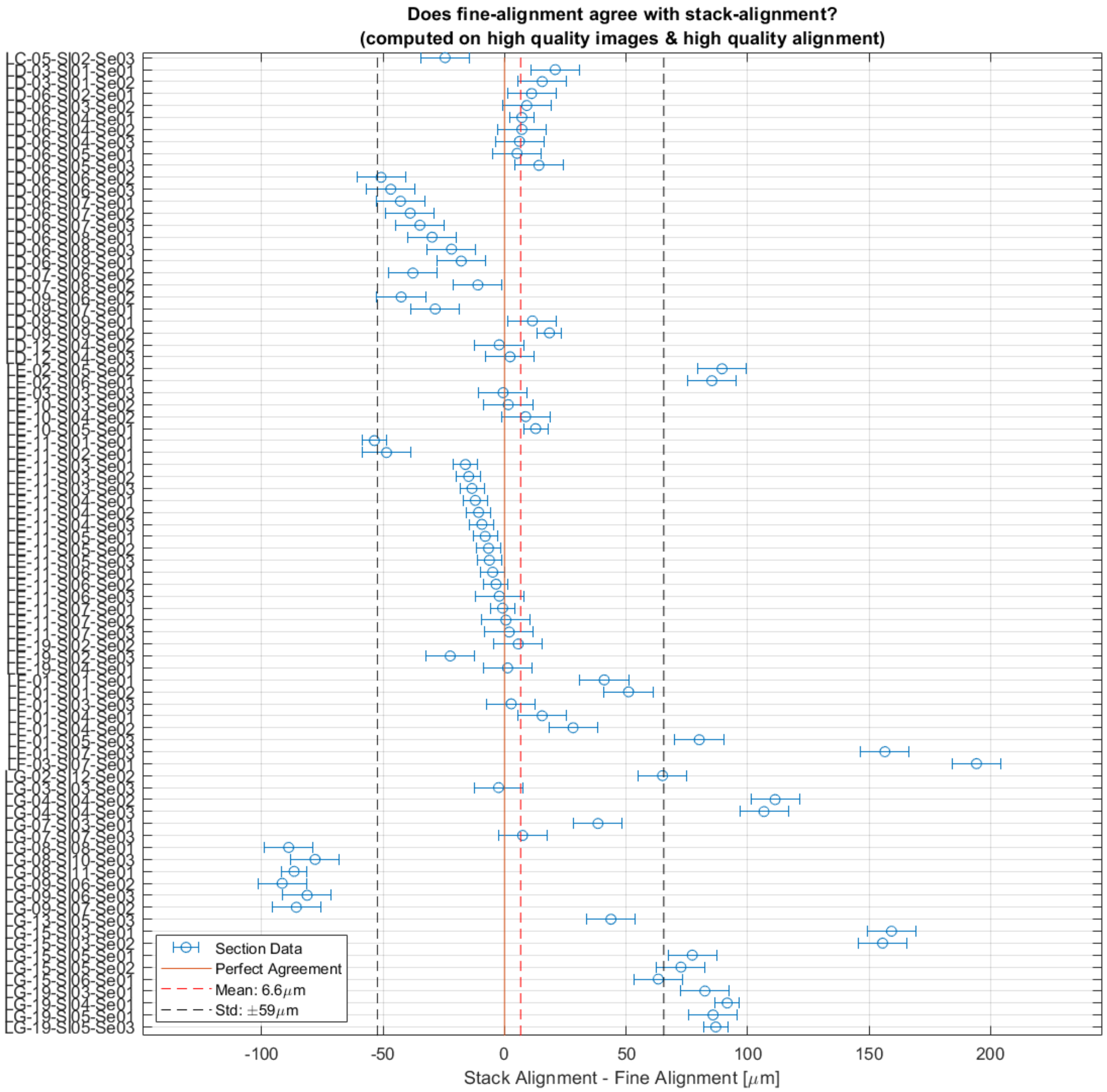


**Figure S9** Stack alignment vs. fine alignment for tissue sections. Each data point on the y-axis is a single tissue section. The x-axis shows the in-plane distance between the stack alignment (that produced by the alignment algorithm) and the fine alignment. Most (one standard deviation) of the stack alignment tissue sections are within 60 microns of the fine alignment. The final precision after fine alignment is shown by the blue error bars, which is the out-of-plane distance the user empirically reports he or she can move before observing a misalignment between the H&E and OCT images. The average precision of the fine alignment is about 25 microns.

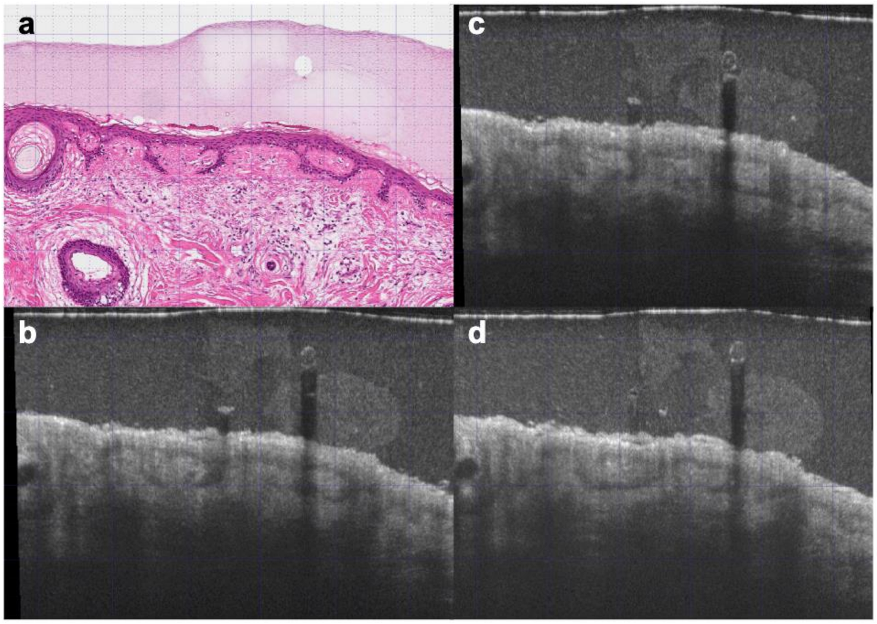


**Figure S10** Image panels illustrate the process of fine alignment on images from human skin from a male, age 66, scalp, obtained by Mohs surgery. a) an H&E stained tissue section and the corresponding fine aligned OCT section b). The effect of shifting the OCT plane in either c) +25 microns or d) -25 microns in the out-of-plane direction. The Rete peg structures in the epithelium are observed to resemble the H&E most closely in b), and 25 micron shifts in either direction lead to apparently worse alignment.

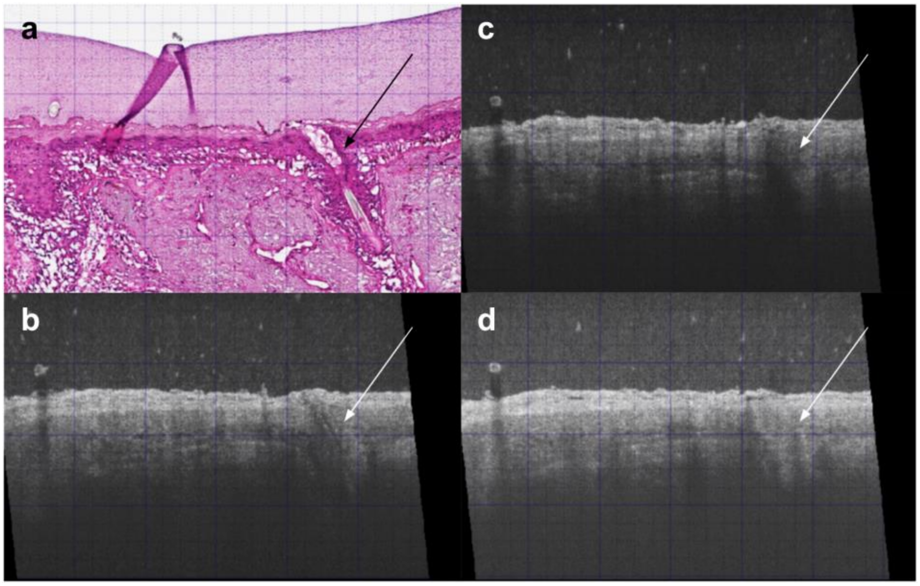


**Figure S11** Image panels illustrate the process of fine alignment on images obtained by Mohs surgery. a) an H&E stained tissue section and the corresponding fine aligned OCT section b). The effect of shifting the OCT plane in either c) +25 microns or d) -25 microns in the out-of-plane direction. The hair follicle pointed to by the arrow is.

### Machine Learning

We tested several general adversarial networks, including pix2pix, pix2pixhd, and cyclegan. Previous work has demonstrated that the pix2pix is less sensitive to nonuniform sample type distribution of the input data, and for these reasons, we used the pix2pix model[2].

We resize both OCT and H&E image to 10x magnification, or 1 micron per pixel, and take crops of size 1024x512. We use the default option for ‘preprocess’ of ‘resize_and_crop’, which resizes all images to 286x286 and then takes a random crop of size 256x256 during training time. The crops thus have magnifications of 2.79x and 5.59x, and pixels sizes of 3.58 microns and 1.79 microns, in the horizontal and vertical directions respectively. The generated OCT images have size 256x256, and are resized back to 1024x512, whereupon they have a magnification of 10x again.

The above method of image generation was found to yield images of superior quality compared to training on 10x image crops of 256x256 with no preprocessing and then testing on larger images of arbitrary size, taking advantage of the fully convolutional nature of the generator. We conjecture that this could be due to improper normalization by the batchnorm function when testing on larger images, as our images contain a large amount of black area. We found that running batchnorm in evaluation mode, using the running mean and variance, also led to suboptimal results.

Examples of generated images are shown in **Figure S12**, with zoom-ins on the epithelium and the entire epithelium segmented in blue.

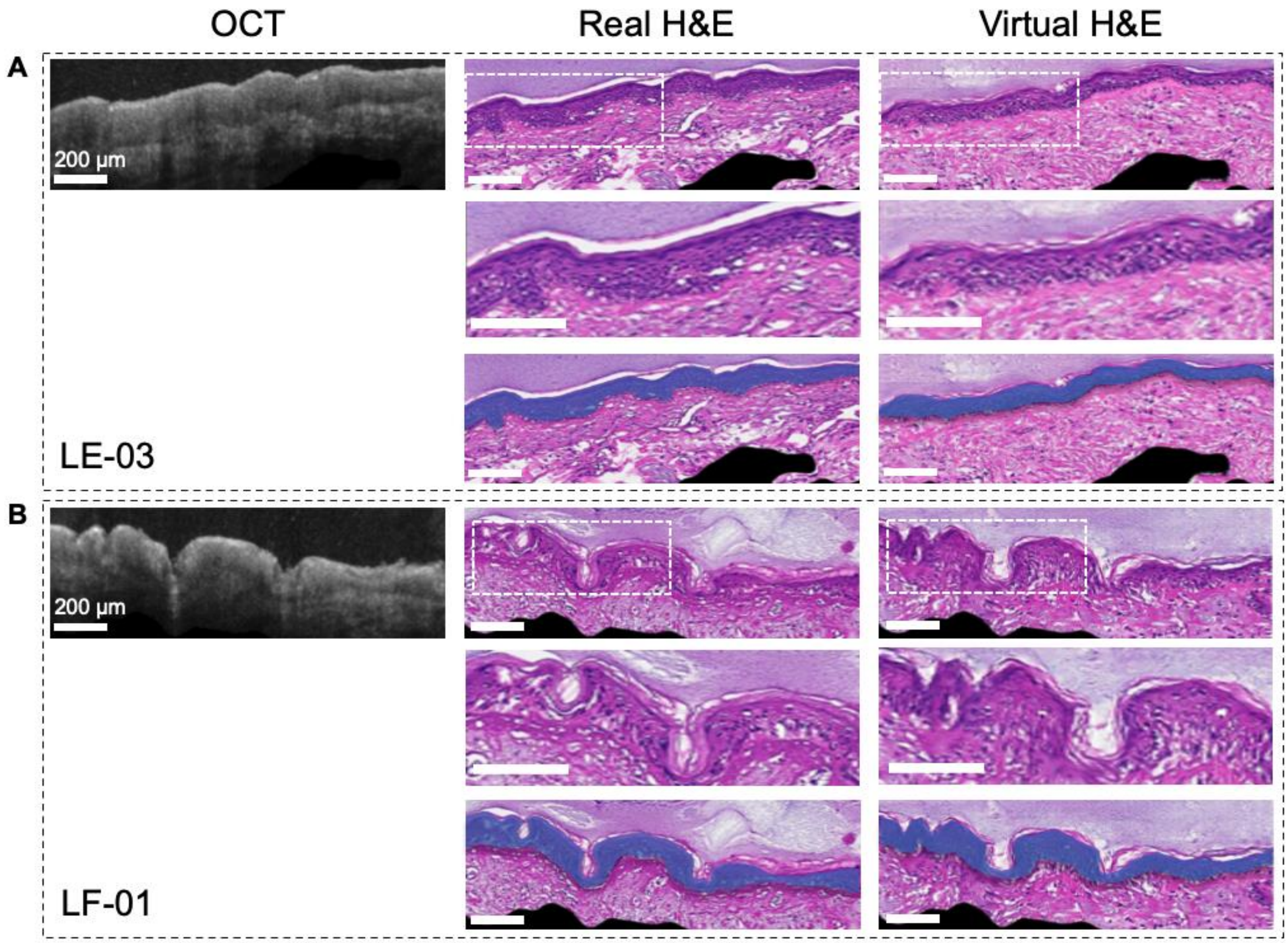


**Figure S12** Further analysis of virtual H&E images where both panels (a) and (b) are skin images from the face. The first row of each panel shows the OCT image, the real H&E image, and the virtual H&E image, from left to right. The second row shows a zoom in of the dotted box in the first row images, for the real H&E image and virtual H&E, from left to right. The last row shows the real and virtual H&E images, from left to right, with the epithelium segmented in blue.

#### How Lack of Alignment Impacts Results

As described above, we tested several general adversarial networks, including Pix2pix[3], Pix2pixHD[4], and CycleGAN[5]. Pix2pix and Pix2pixHD have stringent data requirements for a task to be learned successfully, namely the training examples must be well-aligned[3,4]. In contrast, the CycleGAN model does not require paired training images[5]. We trained a CycleGAN model in order to test how lack of alignment impacts the machine learning results. The model was trained on the same data set as that described in the main text with some organizational differences to ensure the image pools are completely unpaired. The training set was composed of the original OCT test set and the original H&E training set. The trained model was then tested on the OCT test set. Although, nominally, the OCT test set was part of the training data, there were no ground truth H&E labels provided. Thus everything the model had learned about the test images came from unpaired data. We found that the resulting CycleGAN H&E images had a very generic appearance that did not correspond strongly with either the OCT nor real H&E image (**Figure S13**) . The CycleGAN generated epithelium does not match the expected epithelial thickness and the stromal texture was also very different from the ground truth H&E. The results show the importance of using paired images to facilitate learning of how OCT image features correspond to H&E image features. Although the CycleGAN model was able to generate passable H&E sections, it failed in using the OCT image to reproduce important tissue characteristics found in the real H&E image.

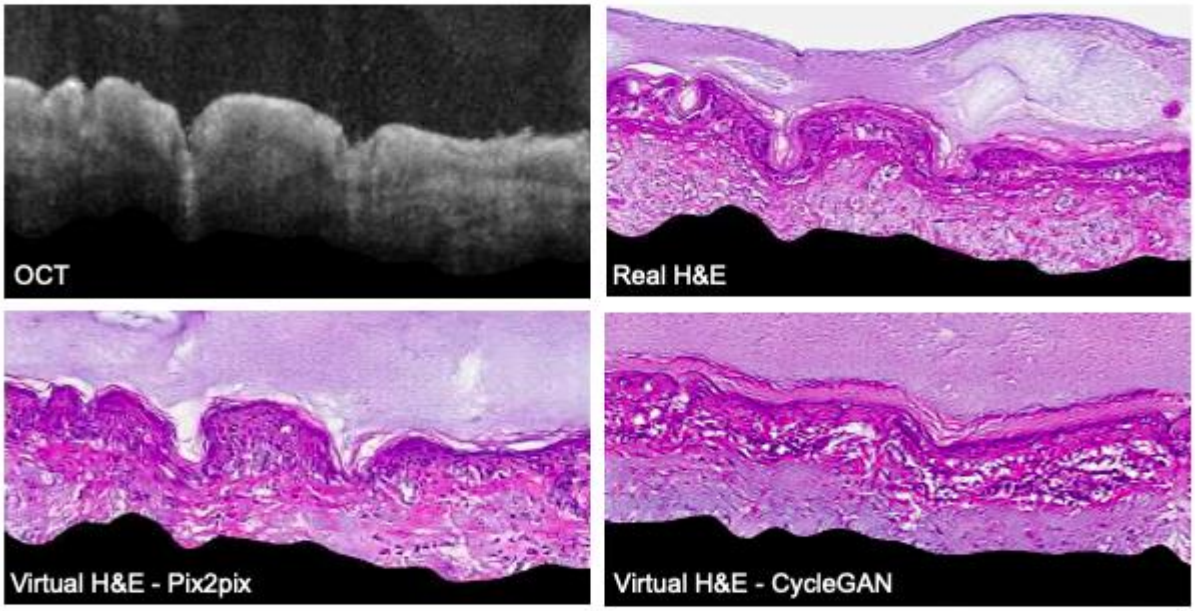


**Figure S13** Virtual histology results from two different adversarial networks; Pix2pix and CycleGAN. Compared to Pix2pix, CycleGAN does not require pair images for training. The CycleGAN model fails to conditionally generate the correct epithelium shape, thickness, and texture. This emphasizes the importance of aligned-paired data for successful learning of tissue features from OCT images.

### Dataset

The following figure, **Figures S14, S15** show some statistics about the samples we collected for training and testing. The majority of samples are from Males, of age 50 to 90 years old, with predominantly Fitzpatrick skin types 1 and 2, which is graded on a scale of 1 to 5, with lighter skin type indicated by lower numbers.

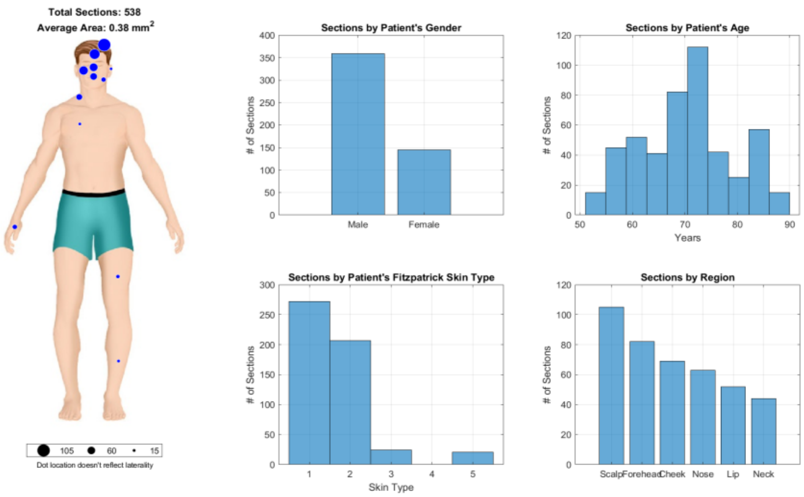


**Figure S14** A plot illustrating where from the skin tissue samples were collected during Mohs surgery. Subjects in the training set. Additional statistics about the gender, age, and Fitzpatrick skin type of patients are shown.

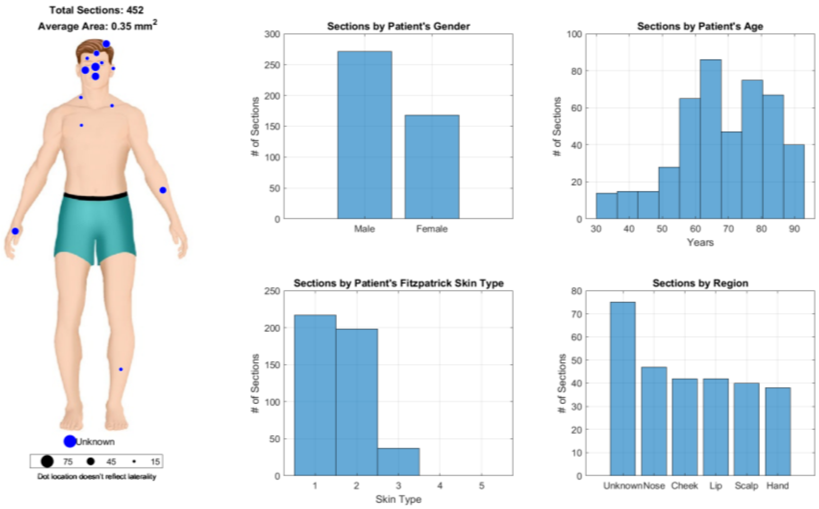

**Figure S15** A plot illustrating where from the skin tissue samples were collected during Mohs surgery. Subjects in the testing set. Additional statistics about the gender, age, and Fitzpatrick skin type of patients are shown.

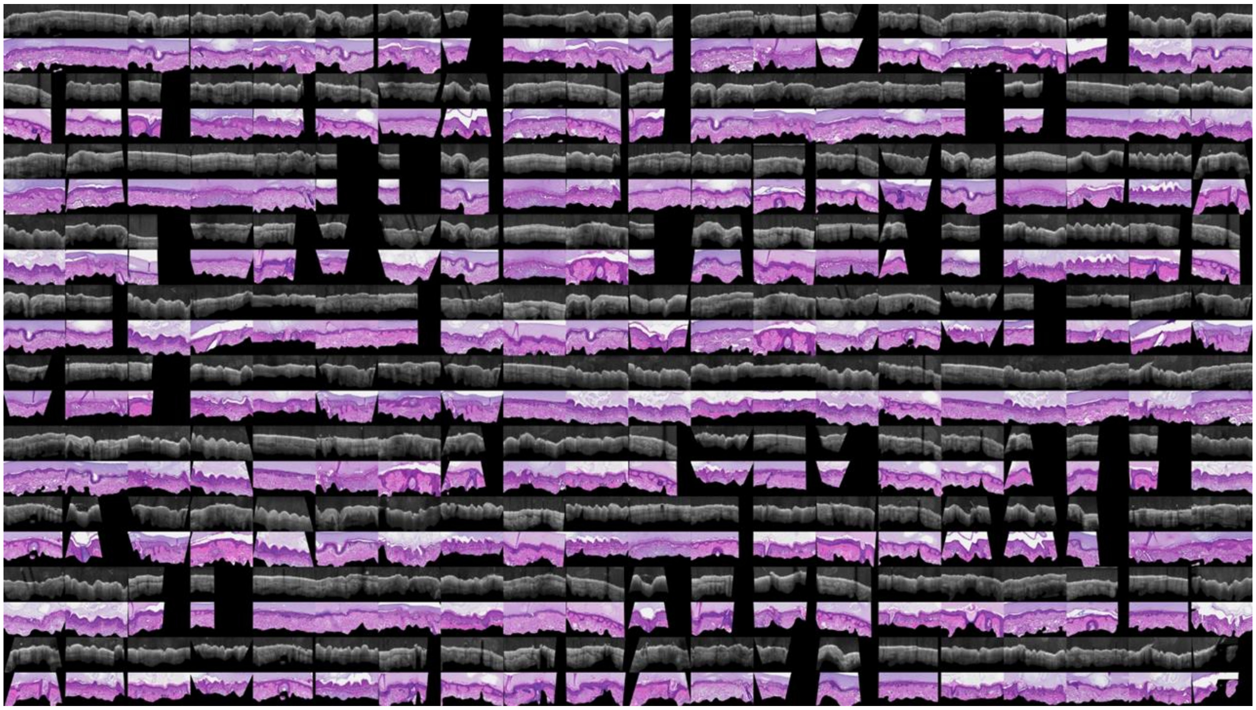

**Figure S16** A compilation of various aligned OCT-H&E image pairs used to train and test the machine learning model (partial list).

### Additional Samples From Test Set

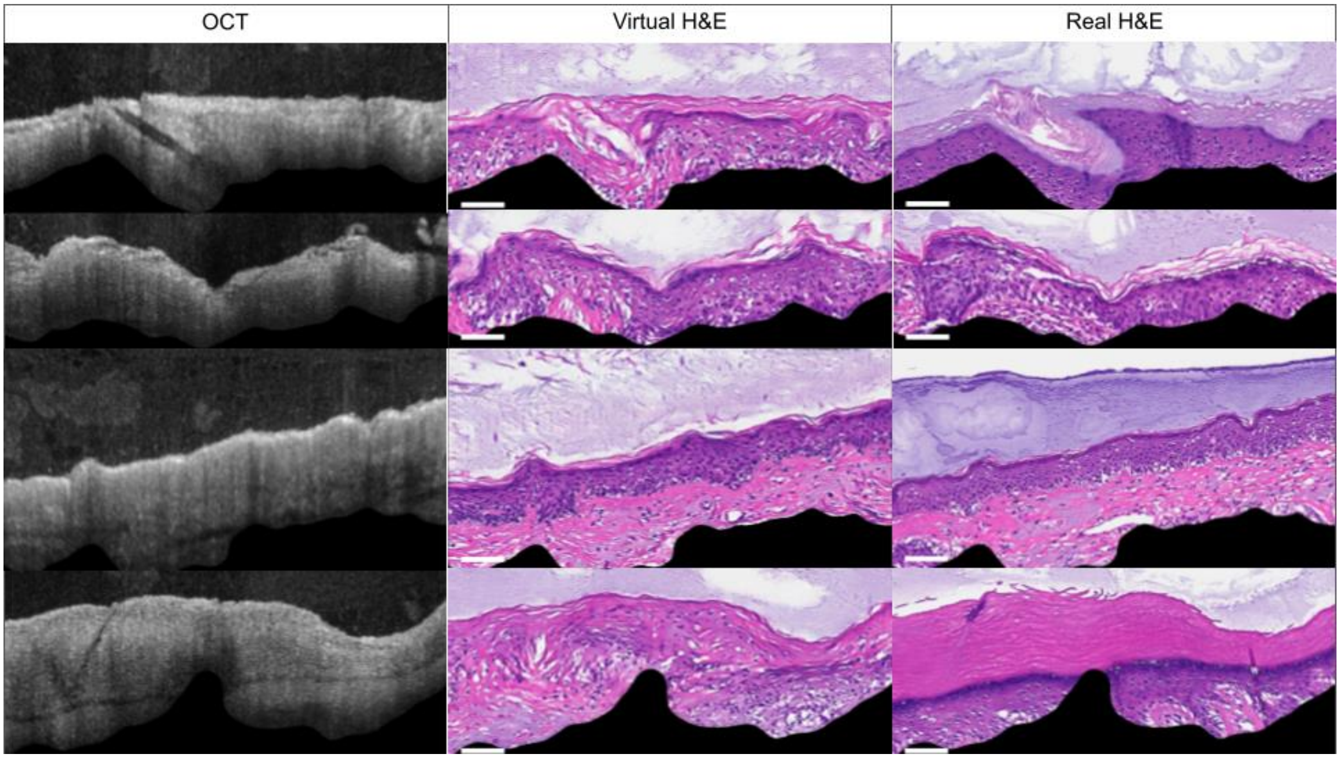


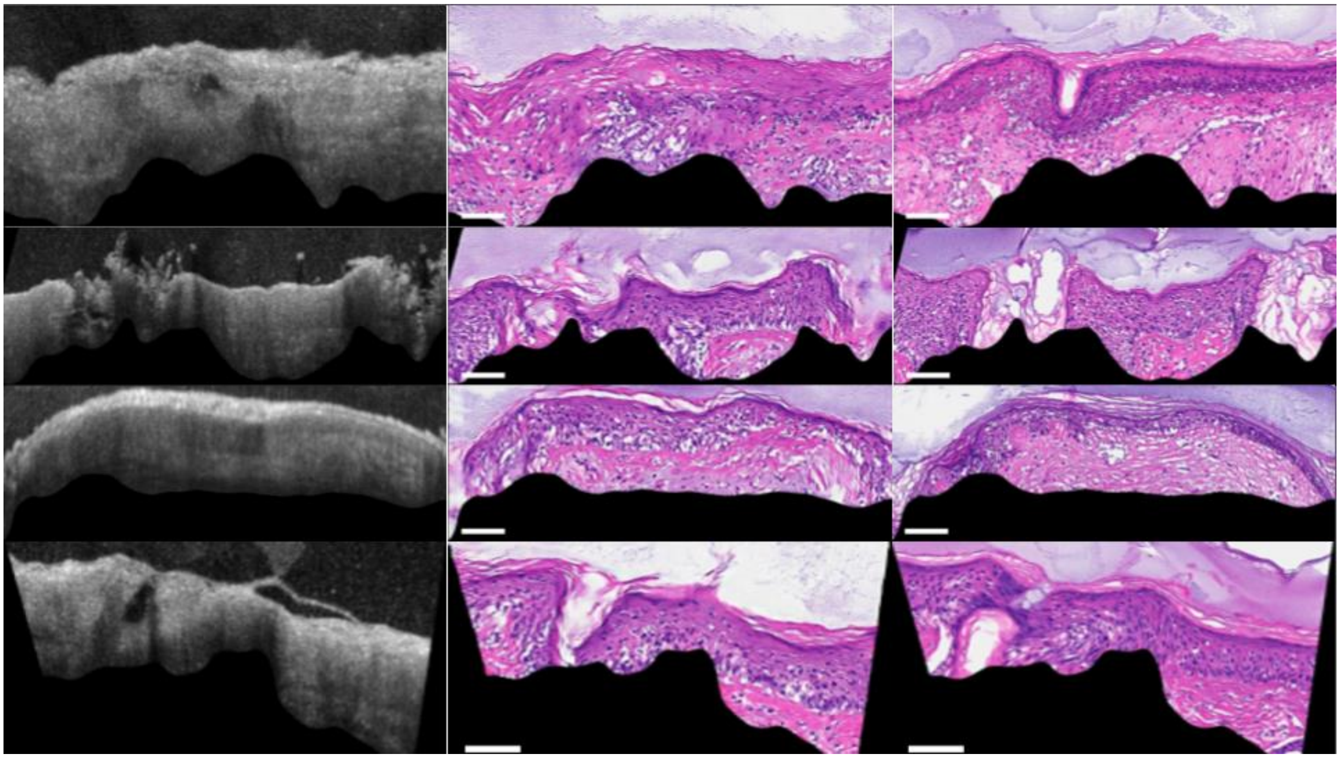

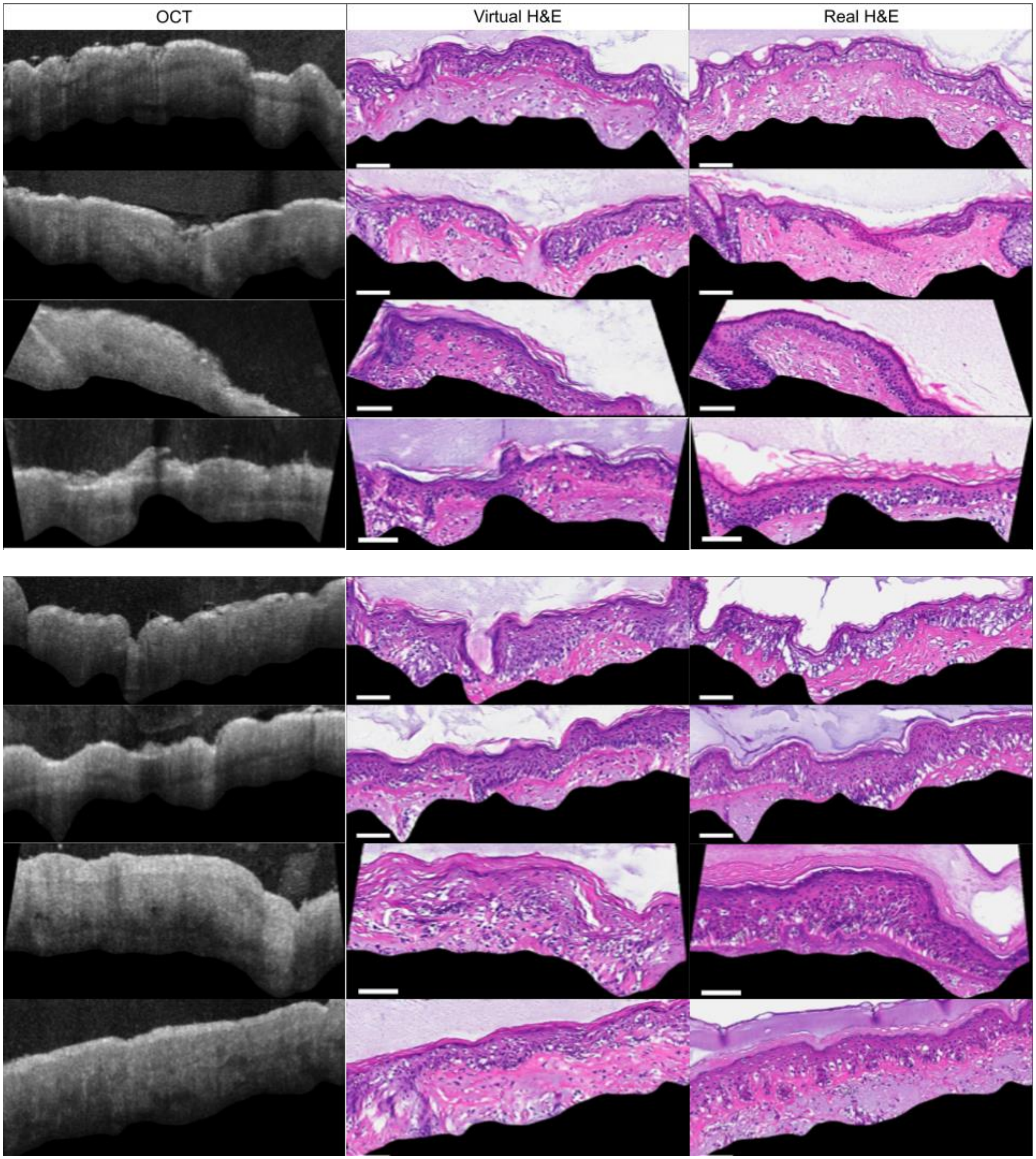


**Figure S17:** Algorithm performances on additional representative examples from the test set, one subject per row. As can be seen, algorithm performance varies depending on the subject. In all of the examples above histology features are present in OCT which support our assumption that increasing the dataset will yield more consistent results. Scale bar is 100 microns.

#### 3D Volume

The 3D H&E volume is obtained by processing an OCT volume slice by slice. The surface of the tissue is segmented from the OCT b-scans by identifying the high intensity tissue-gel interface. All individual segmented slices are interpolated and median filtered before being applied to the generated H&E volume. We observe that the H&E slices obtained when reslicing the OCT volume along the perpendicular direction have a noisy and inconsistent quality, as each column in these images is from a different virtual H&E slice. For presentation purposes, we replace the first and last H&E reslices along the perpendicular direction by two virtual H&E images generated explicitly from the perpendicularly resliced OCT volume.

#### Basal-Cell Carcinoma Preliminary Results

Using the same protocol and methods described in this paper, we scanned and aligned 7 BCC samples. 4 BCC samples (60 sections) were added to the training sets and the model was retrained without any explicit knowledge of BCC present in the image. Promising preliminary results on one subject from the test set is shown below (Fig. S18). We estimate that 50 to 100 additional BCC samples are required to make this algorithm sufficiently robust.

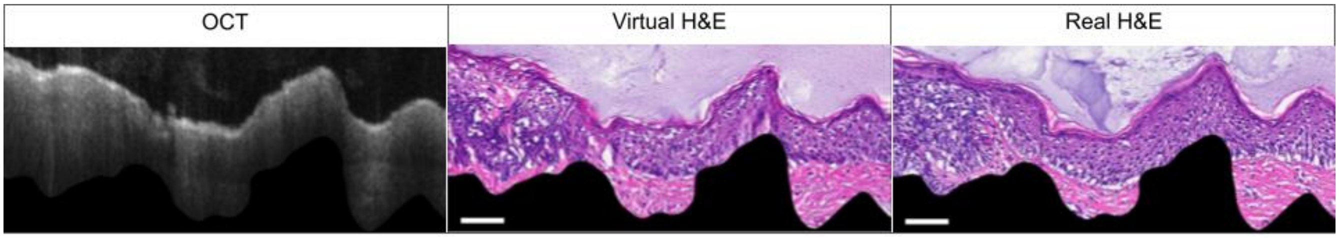

**Figure S18**: Preliminary results show that our OCT2Hist algorithm can generate distinctive cancerous features in the virtual H&E image when adding BCC examples to the train set.
